# Differential task-related fMRI effects of a single dose of guanfacine and lisdexamfetamine in youth with ADHD: a randomised, double-blind, placebo-controlled crossover study

**DOI:** 10.64898/2026.10.01.26364506

**Authors:** Sahid El Masri, Olivia S. Kowalczyk, Marion Criaud, Steve Lukito, Ndabezinhle Mazibuko, Gonzalo Salazar de Pablo, Violeta Perez-Rodriguez, Orsolya Makos, Sheut-Ling Lam, Samuel Westwood, Alexander Eaton-Turner, Natali Bozhilova, Aldo Alberto Conti, Stephanie Stephenson, Maria Rogdaki, Nicoletta Adamo, Lena Johansson, Dennis Ougrin, Fatema Almowasi, Paramala Santosh, Veit Roessner, Gregor Kohls, Mitul A. Mehta, Katya Rubia

## Abstract

**Background:** Stimulant and non-stimulant medications increase fronto-striato-thalamic activation in youth with Attention-Deficit/Hyperactivity Disorder (ADHD). However, comparative effects of the more recently licensed stimulant lisdexamfetamine and non-stimulant guanfacine on functional magnetic resonance imaging (fMRI) correlates in ADHD are unknown.

**Methods:** This exploratory randomised, placebo-controlled, double-blind, crossover study compared fMRI activation in youth with ADHD during a tracking stop-signal task (SST; N = 12), a working memory task (N-back; N = 11) and a sustained attention task (SAT; N = 10), after single clinical doses of either guanfacine extended-release, lisdexamfetamine, or placebo, with one-week washouts. Twenty-one demographically matched typically developing controls (TD) were assessed once to test for potential normalisation of case-control activation differences.

**Results:** Region of interest analysis showed increased right caudate and putamen activation for lisdexamfetamine versus placebo during N-back and SAT, respectively. Both drugs shared an increase in left superior temporal activation versus placebo during N-back. In whole-brain analyses only lisdexamfetamine increased precuneus/posterior cingulate activation during SST versus guanfacine, and during N-back in pre- and postcentral and occipital cortices versus placebo. During SST, placebo-treated ADHD versus TD showed reduced putamen, insula, parietal and occipital activation that was potentially attenuated/normalised under lisdexamfetamine, while under guanfacine lingual gyrus underactivation remained, with additional precuneus and middle temporal underactivation. Also, only lisdexamfetamine improved and normalised SST performance.

**Conclusion:** Lisdexamfetamine appears to have stronger and drug-exclusive upregulation and potential attenuation/normalisation effects than guanfacine on basal ganglia, posterior parieto-occipital and sensorimotor activation during ADHD-relevant executive function tasks with some shared superior temporal and putamen effects.

**Trial registration:** ClinicalTrials.gov (NCT03333668; https://clinicaltrials.gov/study/NCT03333668).

## 1. Introduction

Attention-deficit/hyperactivity disorder (ADHD) is among the most common neurodevelopmental conditions, affecting up to 8 % of children worldwide (Ayano et al., 2023). ADHD is defined by the Diagnostic and Statistical Manual of Mental Disorders (DSM)-5-TR as persistent and developmentally inappropriate symptoms of inattention and/or hyperactivity/impulsivity (American Psychiatric Association, 2022). ADHD is associated with increased likelihood of co-occurring neurodevelopmental and psychiatric conditions, such as oppositional defiant disorder (ODD) followed by specific phobias, conduct disorder (CD), depressive disorders, and autism spectrum disorder (Njardvik et al., 2025), as well as poor social, cognitive and academic outcomes (Faraone et al., 2024). Meta-analyses of functional magnetic resonance imaging (fMRI) studies have shown consistent brain underactivation in youth and adults with ADHD relative to typically developing (TD) age-matched controls in predominantly right-hemispheric dorsolateral prefrontal, parietal and striato-thalamic regions during attention tasks (Hart et al., 2013), in right inferior frontal/insula and striatal regions during cognitive control tasks (Hart et al., 2013; Lukito et al., 2020; McCarthy et al., 2014; Norman et al., 2016), in bilateral dorsolateral prefrontal regions during N-back working memory (WM) tasks (McCarthy et al., 2014) and in inferior frontal, parietal and cerebellar regions during timing tasks (Hart et al., 2012). Recent cross-task meta-analyses confirm differences in left and right inferior, dorsolateral prefrontal and striatal regions with more pronounced differences in youth compared to adults (Tamon et al., 2024; Yu et al., 2023).

There is evidence for delayed functional maturation of resting-state networks, especially in ventral and dorsal attention and cognitive control networks and the default mode network (DMN). Furthermore, there is evidence for poorer anticorrelation between these networks and the DMN which has been associated with mind-wandering interference (Sripada et al., 2014). Structural mega-analyses (Hoogman et al., 2017, 2019) and a meta-analysis (Yu et al., 2023) have reported cortical thickness and surface-area differences in frontal and temporal regions, volumetric differences in basal ganglia (BG), limbic, and thalamic regions, and structural differences in the corpus callosum. Moreover, there is evidence for neurotransmitter differences, most prominently in dopaminergic and noradrenergic systems (Faraone, 2018).

First-line pharmacological treatment for ADHD generally involves stimulant medication, although the recommended agent and treatment sequence vary by age and national guidelines. Commonly used stimulants include methylphenidate, (dex)amphetamine and lisdexamfetamine, which show moderate-to-large effect sizes in reducing ADHD symptoms (Gosling et al., 2025), via inhibition of dopamine and norepinephrine transporters in the brain, and, in the case of amphetamines, additional promotion of monoamine release (Faraone, 2018). Non-stimulant medications are commonly considered when stimulants are ineffective, not tolerated, contraindicated, or not preferred, such as atomoxetine, viloxazine, bupropion, guanfacine and clonidine, which all share some noradrenergic or adrenergic mechanism (Fu et al., 2022; Neuchat et al., 2023; Verbeeck et al., 2017; Yu et al., 2020); atomoxetine is associated with moderate reductions in ADHD symptoms in both children/adolescents and adults (Cortese et al., 2018; Gosling et al., 2025; Ostinelli et al., 2025).

The neurofunctional mechanisms of action of these medications are relatively poorly understood, particularly for more recently licensed stimulants and non-stimulants. Our meta-analysis of 14 fMRI datasets on single-dose stimulants during executive function and timing tasks in youth with ADHD showed that stimulants relative to placebo or off medication increased activation of the right inferior frontal cortex (IFC), followed by BG at a more lenient threshold and led to reduced activation in mid-frontal areas of the default mode network (DMN) (Rubia et al., 2014), which are associated with mind-wandering (Menon, 2023).

A more recent meta-analysis of studies in youth and adults with ADHD testing the effects of stimulants (19 studies) and non-stimulants (nine studies) on fMRI activation during tasks of cognitive control and reward processing, combining pre–post treatment, medication-versus-placebo/control, and group-by-time contrasts (Pan et al., 2024) showed converging increases in activation in the left supplementary motor area (SMA) across these medication-effect contrasts, suggesting possible normalisation towards TD activation patterns. Further seen were increased activation in left superior frontal, mid-cingulate cortices and amygdala under stimulants relative to non-stimulants, and reduced activation of left prefrontal and right caudate regions under non-stimulants relative to control groups.

These meta-analytic findings (Pan et al., 2024; Rubia et al., 2014) align well with our single-study fMRI findings of differential methylphenidate and atomoxetine effects on brain activation in youth with ADHD (Cubillo et al., 2014a, 2014b; Kowalczyk et al., 2019; Smith et al., 2013). Specifically, during inhibitory control tasks, both methylphenidate and atomoxetine normalised left VLPFC underactivation, while atomoxetine enhanced right inferior parietal/superior temporal activation relative to placebo; only methylphenidate enhanced right VLPFC/STG activation relative to atomoxetine and placebo and showed significant normalisation of cerebellar underactivation (Cubillo et al., 2014b). During WM, atomoxetine increased activation in the right dorsolateral prefrontal cortex (DLPFC) compared to methylphenidate, whereas methylphenidate increased the left IFC activation relative to placebo and atomoxetine, suggesting drug-specific laterality effects, while both drugs enhanced fronto-temporo-striatal activation relative to controls (Cubillo et al., 2014a). Activation in the left IFC and insula was also increased during WM after a single dose of methylphenidate compared to no drug in youth with ADHD (Prehn-Kristensen et al., 2011). Additionally, both methylphenidate and atomoxetine normalised activation differences in youth with ADHD under placebo in parietal, temporal, precuneus/posterior cingulate, and striato-insular regions during sustained attention with drug-selective activation of left ventral fronto-temporal regions for methylphenidate relative to placebo and atomoxetine (Kowalczyk et al., 2019), and both drugs normalised underactivations in right VLPFC/insula regions in youth with ADHD during timing tasks (Smith et al., 2013).

Few studies tested the effects of the more recently licensed ADHD drugs, lisdexamfetamine or guanfacine, on task-related fMRI brain activation, and none directly compared their effects in the same individuals. A network meta-analysis examining efficacy and tolerability of several ADHD drugs showed higher efficacy for amphetamines, including lisdexamfetamine, than for methylphenidate in improving symptoms in both youth and adults with ADHD and supported amphetamines as first-line treatment for adults with ADHD, but methylphenidate for youth when efficacy and safety were considered together (Cortese et al., 2018). So far, only two fMRI studies have tested the effects of lisdexamfetamine (Schulz et al., 2018) and guanfacine (Bédard et al., 2015) during executive functioning, i.e. Go/No-Go tasks in individuals with ADHD, although the task in the study on lisdexamfetamine used an emotional Go/No-Go paradigm that includes emotionally salient stimuli (Schulz et al., 2018), generally considered a measure of “hot” executive function versus emotionally neutral “cool” executive functioning tasks. In 25 youth with ADHD, 6–8 weeks of guanfacine treatment did not produce a significant overall effect on response-inhibition-related activation during a Go/No-Go task; however, compared with placebo, greater clinical improvement under guanfacine was associated with greater pre-to-post reductions in the right midcingulate cortex (MCC)/supplementary motor area (SMA) and left posterior cingulate cortex (PCC) (Bédard et al., 2015). Similarly, 3–4 weeks of lisdexamfetamine treatment in 25 adults with ADHD increased right amygdala activation during responses to sad faces compared with placebo, with the relative increase correlating with symptom improvement (Schulz et al., 2018). Furthermore, a study using functional near-infrared spectroscopy (fNIRS) found no effect of a single dose of guanfacine on brain function in 12 youth with ADHD during a non-emotional/neutral Go/No-Go task (Ikeda et al., 2021).

Given the sparse and inconsistent brain function findings of these more recently licensed ADHD drugs, we compared the differential effects of single doses of lisdexamfetamine and guanfacine extended-release relative to placebo during three typically compromised fMRI tasks in ADHD, including a sustained attention task (SAT), a tracking stop-signal task (SST) and a WM task (N-back task) in youth with ADHD using a randomised, placebo-controlled, double-blind, crossover design. TD were assessed once to test for potential normalisation of case-control activation differences.

Given meta-analytic evidence that methylphenidate increases right IFC/insula and BG activation relative to placebo (Rubia et al., 2014), and evidence for drug-selective effects of methylphenidate over atomoxetine and placebo on left IFC during SAT (Kowalczyk et al., 2019), WM (Cubillo et al., 2014a), and on right fronto-striato-cerebellar activation during response inhibition (Cubillo et al., 2014b), we hypothesised that lisdexamfetamine would show drug-specific effects on IFC and insular-striatal regions during those three tasks compared to guanfacine. For the N-back task, based on evidence for drug-specific right DLPFC enhancement with atomoxetine over methylphenidate (Cubillo et al., 2014a), we hypothesised that guanfacine would increase activation in the right DLPFC in a drug-selective manner compared to lisdexamfetamine and placebo. Although alpha-2 agonists like guanfacine differ from noradrenergic reuptake inhibitors like atomoxetine at the molecular level, they nevertheless act on noradrenergic/adrenergic prefrontal mechanisms implicated in arousal, cognition, and ADHD symptom regulation (Neuchat et al., 2023). For the case-control comparisons, we hypothesised that lisdexamfetamine compared to guanfacine would have a clearer effect of normalisation of task-relevant underactivations in youth with ADHD under placebo.

## 2. Methods

This double-blind, placebo-controlled, randomised study was registered on ClinicalTrials.gov (Identifier: NCT03333668) before data collection commenced: https://clinicaltrials.gov/study/NCT03333668

### 2.1. Determination of sample size

The prespecified target sample size was 20 participants with complete ADHD datasets, as documented in the study protocol. This target was consistent with previous methodological recommendations suggesting a minimum sample size of 20 for fMRI comparisons (Thirion et al., 2007) and was further informed by a power analysis based on a previous within-subject fMRI study comparing the acute effects of atomoxetine and methylphenidate using the same working-memory task (Cubillo et al., 2014a). To obtain 20 complete datasets, the protocol planned to screen 50 patients to account for ineligibility and attrition.

### 2.2. Participants

Twenty-five youth aged 8–20 years meeting clinical diagnostic criteria for predominantly inattentive, predominantly hyperactive/impulsive, or combined presentation of ADHD based on the DSM-5 criteria were recruited from NHS Trusts, private clinics in the Greater London area, social media, ADHD-and parent-support groups, and from the database of another study between 2019 and 2024; five dropped out after at least one of the three fMRI visits, due to an adverse event (dizziness, light-headedness, trouble keeping their eyes open and staying upright; n = 1), discomfort from scanner noise (n = 2), excessive head tilt and motion during the first fMRI visit (n = 1), or school/university workload-related stress (n = 1). Participants were excluded from a given task contrast analysis if more than 20 % of volumes in the corresponding acquisition met the criterion for a motion outlier. Section 2.6 details the motion metrics and thresholds. After the exclusion criteria were applied to the 20 participants who completed the study, six participants were excluded across all task analyses due to being a motion outlier (N = 5) and because of missing structural MRI data due to data loss during the post-scan transfer/upload process to the university’s internal data platform (N = 1), leaving the final sample with 14 participants (mean age = 15.2, SD = 2.4). However, the final task-specific samples further ranged between 10 and 12 participants: four participants from the SAT, three participants from the N-back task and one participant from the SST were excluded because they were motion outliers, as well as an additional one in the SST, who was excluded from the successful stop vs go trial contrast but who had enough frames unaffected by motion for the failed vs go trials contrast. Lastly, one more participant was excluded only from the failed vs go trials contrast, but not from the successful stop vs go trial contrast, because the proportion of successful inhibitions exceeded the typical adaptive tracking range (> 80%). The final sample sizes for each task are therefore 10 participants for the SAT, 11 participants for the N-back task and 12 participants for each contrast of the tracking SST (with 11 participants overlapping and one participant unique to each SST contrast). Participant flow through the study, with reasons for drop-out, is shown in the Consolidated Standards of Reporting Trials (CONSORT) flow chart (see Figure 1). It should be noted that counts are based on the participant records that remained available. Additional initial contacts may have occurred via the study’s Facebook account, but these could not be verified because the account and its message history were no longer accessible. CONSORT reporting guidelines were followed (Schulz et al., 2010). The completed CONSORT checklist for randomized crossover trials is provided in Supplementary data 6 (Dwan et al., 2019).

**Figure 1.**
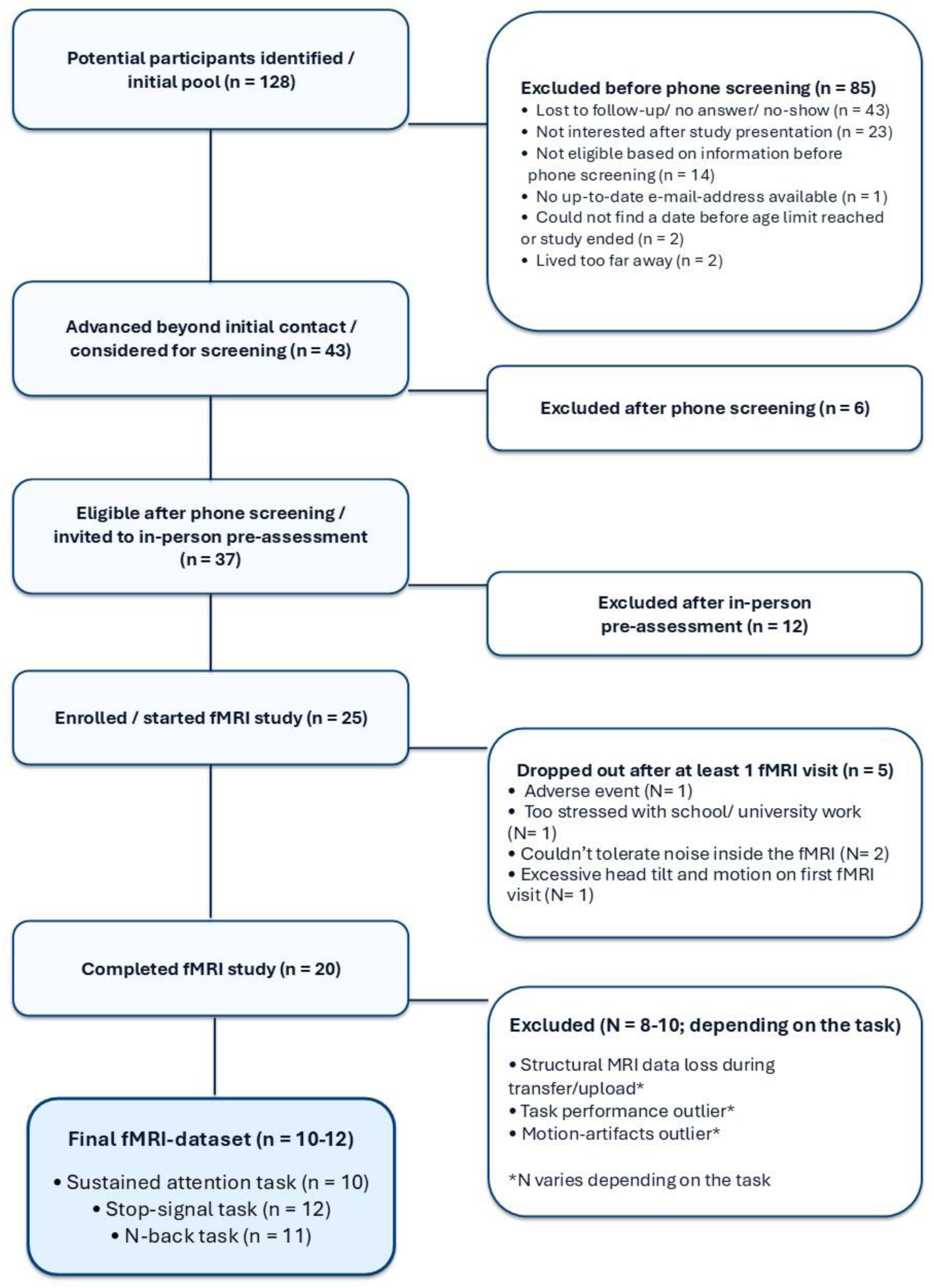
Consolidated Standards of Reporting Trials (CONSORT) flow chart.

Participants with ADHD were medication-naïve (N = 4) or stopped taking stimulants for at least 48 hours before each of the three testing sessions or for at least two weeks for any other psychoactive medications. Participants completed an in-person pre-assessment, including administration of an electrocardiogram, measurements of blood pressure, body weight, Edinburgh Handedness Inventory (Oldfield, 1971), IQ using the WASI-II (Wechsler, 2011) and a mock MRI scanner visit. Inclusion required a score above clinical cut-offs in the ADHD Rating Scale-IV (ADHD-RS; (DuPaul et al., 1998), total score ≥24, approximately the 90th percentile), Conners 3rd Edition–Parent (Conners 3-P; (Conners, 2008), DSM-IV-TR criteria: 6 or more symptoms for either inattention or hyperactivity or both) and the Kiddie Schedule for

Affective Disorders and Schizophrenia (K-SADS; (Kaufman et al., 1997, 2016); DSM-5 criteria: six or more symptoms for either DSM-5 inattention or hyperactivity or both). Participants were excluded if their WASI-II Full Scale IQ–4 (FSIQ-4) was below 70 or if they had MRI contraindications, a history of substance abuse, a neurological condition or any other major clinical psychiatric disorder except for mild mood and depression symptoms, autism, phobias, anxiety, conduct disorder, oppositional defiant disorder, or eating disorder. We also excluded participants with learning disabilities, reading, speech, or language disorders or other somatic illnesses that would impact data integrity or safety of the participant (i.e. medication contraindications, cardiovascular problems).

Twenty-four participants in the TD group were matched for sex, age, and IQ with the ADHD group, of whom 21 (mean age = 14.9, SD = 3.2) were retained after 3 participants were excluded because they had outlier values in a minimum of three performance-related measures (minimum 1.5 interquartile ranges). For one additional TD participant, SAT and N-back task data were unavailable; the reasons were not documented in the available study records. Consequently, the TD group comprised 21 participants for the SST and 20 participants for the SAT and N-back task. Participants in the TD group were assessed once without medication to examine potential normalisation effects of the two drugs on fMRI activation and cognitive differences compared to participants with ADHD under placebo. Inclusion criteria for the TD participants were an IQ above 70, a score below the cut-off (i.e., 13) for autistic traits on the Social Communication Questionnaire (SCQ) (Rutter et al., 2003) and no more than five symptoms in each of the Conners 3-P DSM-IV-TR inattention and hyperactivity/impulsivity domains and no history of substance abuse, neurological deficits, MRI contraindications, learning disabilities, reading, speech or language disorder or other illness that would impact data integrity or safety of the subject.

Written informed consent was obtained from participants aged 16 years or older. For participants younger than 16 years, written informed consent was obtained from a parent or legal guardian, and written assent was obtained from the participant. Participants received £20 for the pre-assessment visit and £50 for each testing/scanning session. Ethical approval was obtained from the London Bridge Research Ethics Committee (REC; 18/LO/0472). The study was conducted in accordance with the Declaration of Helsinki.

### 2.3. Testing procedures

Participants with ADHD made one in-person visit for pre-assessments followed by three fMRI visits ≥7 days apart. Guanfacine extended-release has a half-life of approximately 16 hours (FDA, 2019), while the active metabolite d-amphetamine after lisdexamfetamine administration has a half-life of approximately 9–15 h (Ermer et al., 2016). Seven days was considered a sufficient washout period between visits to avoid drug interactions based on established guidelines of a minimum washout of 8.5 half-lives (Chow, 2014). On each visit, participants with ADHD received either placebo or a single, typical clinical oral dose of lisdexamfetamine or guanfacine. KR generated and retained the computer-based randomisation list determining the order of the three treatment conditions, balanced equally across the six possible treatment sequences. Participants were enrolled by SEM and other study researchers, none of whom had access to the randomisation list during recruitment or data collection. Maudsley Pharmacy prepared and dispensed the study medications in matching over-encapsulated capsules according to the randomisation sequence. SEM administered the coded medication under clinical supervision while remaining blinded to the treatment condition administered at each visit. Participants underwent fMRI approximately 4.5 hours after medication administration. Two participants experienced transient mild adverse events after lisdexamfetamine, one of whom experienced irritability, dizziness, light-headedness, trouble keeping their eyes open, and difficulty staying upright and ended the study prematurely, whereas the other continued and completed the study. Thereafter, the original doses of lisdexamfetamine (30 mg for participants under 30 kg of body weight and 50 mg for participants over 30 kg of body weight) were adjusted: 8- and 9-year-old participants with ADHD received 20 mg regardless of weight, and participants aged 10–20 years received 30 mg or, if weighing more than 60 kg, 0.5 mg/kg, rounded down to the nearest available dose of 30, 40, or 50 mg. Guanfacine was given at 0.05 mg/kg and rounded down to the nearest 1, 2, or 3 mg. Placebo contained 10 mg vitamin C.

After each of the three fMRI/testing visits, adverse effects (AEs) were assessed using a parent-report questionnaire (Hill, 2001) in adapted form. Compared with the 17 items described in subsequent studies, the version used included ten additional AE items and employed a four-point scale assessing new onset or worsening following treatment, rather than the five-point symptom-frequency scale reported for the 17-item version. For each medication condition, responses across the 27 items were combined into a continuous total AE score, with higher scores indicating a greater overall burden of newly occurring or worsening AEs. The AE analysis included the 14 participants with complete data across all three medication conditions.

### 2.4. fMRI tasks

For schematic figures and more detailed descriptions of the three tasks see Supplementary data 1–3. Sustained attention was assessed with a well-validated modified psychomotor vigilance sustained attention fMRI task (SAT, 12 min) following established protocols (Christakou et al., 2013; Hart et al., 2017; Kowalczyk et al., 2019; Norman et al., 2017). The SAT requires participants to respond quickly to a visual timer after either short, predictable delays (0.5 s, demanding sensorimotor synchronisation) or long, infrequent, unpredictable delays (2, 5, 8 s) that impose a greater sustained attention load. The main contrast compared brain activity during 8-s delay trials with 0.5-s delay trials to differentiate sustained attention demands from anticipatory sensorimotor processes.

Response inhibition was measured with an established and well-validated mixed-trials, event-related and individually adjusted tracking SST (6 min) (Cubillo et al., 2014b; Hart et al., 2014; Rubia et al., 2010, 2013). The SST measures response inhibition with ∼80 % go trials requiring a left or right button press and ∼20 % stop trials requiring inhibition of the motor response. The task dynamically adjusts stop-signal delays to maintain an approximate 50 % successful inhibition rate. The key contrasts were successful stop versus go trials, indexing response inhibition, and failed stop versus go trials, specifically indexing error monitoring associated with unsuccessful inhibition.

The N-back task (6 min 12 s) was adapted from previous block-design paradigms to include only 0-back, 1-back, and 2-back conditions (Chantiluke et al., 2015; Cubillo et al., 2014a; Kobel et al., 2009), excluding the 3-back to ensure a suitable level of difficulty for younger children. Participants respond to letters, indicating whether the current stimulus matches “X” (0-back) or the letter displayed one (1-back) or two trials before (2-back), requiring storage and updating in WM. The contrast used in the analysis was the 2-back > 0-back comparison to index increasing WM load.

### 2.5. fMRI acquisition

MRI data were acquired on a 3T Discovery MR750 MRI scanner (GE Healthcare, Chicago, USA) at the Centre for Neuroimaging Sciences, Institute of Psychiatry, Psychology and Neuroscience, King’s College London, UK. The first two participants with ADHD were scanned using an 8-channel head coil (HDx 8HR Brain), and in the remaining scanning sessions a newer Head Neck Spine coil with 12 channels was used. A high-resolution T1-weighted 3D structural scan (MPRAGE/BRAVO) was acquired (TR = 7.312 ms, TE = 3.016 ms, flip angle = 11°), with a voxel size of 1.0547 × 1.0547 × 1.2 mm, 161 slices (196 × 256 × 161 voxels), no slice gap. T2*-weighted gradient-echo EPI was used (TE = 30 ms; SAT/SST/N-back: TR = 1.5/1.8/2.0 s, in-plane resolution = 3.75/3.30/3.75 mm, and slice thickness/gap = 3.5/1.0, 3.0/0.3, and 3.0/0.3 mm). The full fMRI acquisition parameters for all three tasks are provided in the Supplementary data 4.

### 2.6. Data analysis

All fMRI data were pre-processed (realigned, resliced, coregistered, segmented, spatially normalised and smoothed) using SPM12 (Penny et al., 2007); version 7771 (https://github.com/spm/spm12/releases/tag/r7771). Segmentation was done in the SPM12 toolbox CAT12 (Gaser et al., 2024). Analysis (first and second level) was done using SPM26. Slice-timing correction was not applied. Instead, each condition was modelled using the canonical haemodynamic response function and its temporal derivative to accommodate small variations in response latency.

Motion-outlier volumes were defined using a ≥1-mm threshold for the Euclidean magnitude of frame-to-frame translational displacement. Acquisitions were excluded when >20% of volumes met this criterion. Absolute translation >3 mm and excessive rotation were additionally reviewed as quality-control indicators. Motion-outlier volumes were censored in the first-level model using one separate nuisance regressor per flagged volume. Mean event-related motion impact (flagged volumes that overlapped with task events) was low, ranging from 0.33 % to 2.83 % across task/drug conditions. All volumes from retained acquisitions were included in the first-level models, together with Friston-24 motion regressors and one separate nuisance regressor per flagged volume.

Three separate first-level general linear models were estimated, one for each task, yielding four primary contrasts: 8-s delay > 0.5-s delay (SAT), successful stop > go and failed stop > go (SST), and 2-back > 0-back (N-back). Each contrast was then analysed in a separate second-level flexible factorial model. The models included Subject as a factor accounting for repeated measures, Medication (placebo, lisdexamfetamine, guanfacine) and session order (first, second, or third treatment session) as within-subject factors. All six possible treatment sequences were represented among the 14 participants retained after general fMRI exclusions, with 2, 2, 3, 3, 2, and 2 participants assigned to sequence orders 1–6, respectively. Tested contrasts comprised the main effect of Medication (omnibus F-test) and pairwise medication comparisons: prespecified, hypothesis-informed directional medication contrasts were lisdexamfetamine > placebo, lisdexamfetamine > guanfacine, guanfacine > placebo, and, for the N-back DLPFC hypothesis, guanfacine > lisdexamfetamine and guanfacine > placebo. Second-level analysis was carried out at the whole-brain level and within predefined ROIs. ROI masks were derived from the Automated Anatomical Labelling 1-mm AAL3v2 atlas, April 2024 (Rolls et al., 2020) and saved as individual NIfTI files.

In line with our a priori hypotheses, we selected the following ROIs for each task: bilateral IFC, caudate and putamen for the SAT, bilateral IFC, insula, caudate and putamen for the tracking SST and bilateral DLPFC, IFC, caudate, putamen and superior temporal gyrus (STG) for the N-back task.

ROI analyses used peak-level small-volume correction with family-wise error control within each predefined mask (*p*_FWE,SVC_ < 0.05). No additional multiplicity correction was applied across ROIs; ROI findings should therefore be interpreted as exploratory.

Where the whole-brain omnibus F-test (*p*_FWE_ < 0.05) with a cluster-defining threshold of *p*_unc_ < 0.001 identified at least one significant cluster, subsequent exploratory, hypothesis-informed medication t-contrasts were examined across the whole brain and were not spatially restricted to clusters identified by the omnibus F-test. Given the modest sample size, we adopted an additional conservative reporting criterion and did not report whole-brain clusters comprising fewer than 10 contiguous voxels. Because cluster extent was not part of the inferential criterion in the ROI-analysis, ROI findings with fewer than 10 contiguous voxels remained statistically significant at the peak level but were not emphasised/interpreted in the main text and were transparently indicated in the relevant table/figure legends.

Additionally, medication-specific between-group comparisons were conducted between ADHD participants under each medication condition and TD participants for all main task contrasts using two-sample t-tests in both directions (TD > ADHD and ADHD > TD), at the whole-brain level and within predefined ROIs.

Anatomical labelling was performed using the Harvard–Oxford probabilistic atlas distributed with FSL (FMRIB Software Library, version 6.0.7), queried in MNI152 standard space using FSLeyes. Labels were assigned based on the maximum probability map at each peak coordinate. When the global maximum was located in white matter or was not anatomically informative according to the Harvard–Oxford atlas, the highest local maximum within grey matter was reported. Brodmann areas (BA), where applicable, were identified separately at the same peak coordinates using the Talairach Daemon Labels implemented in FSLeyes. For confirmation of labels and examination of potential cluster extensions, or if Talairach Daemon Labels could not provide a number for BA, whole-brain images were visually inspected. Directional qualifiers were reported for descriptive use in brackets.

Since coil differences can systematically affect signal-to-noise ratio across all scans, we included a covariate of no interest in the analysis of ADHD vs TD, coding the two ADHD participants scanned with the 8-channel coil as 1 and all other participants as 0. For the SAT, however, only one participant scanned with the 8-channel head coil was included. Therefore, coil type was not included as a covariate, because a group represented by a single participant would not allow a reliable estimate of a general coil effect.

Task performance and AEs were analysed in IBM SPSS Statistics (IBM Corp., 2021); version 28.0.1.1. The total adverse-effect score was compared across the three medication conditions using a Friedman test, followed by pairwise Wilcoxon signed-rank tests. The three pairwise adverse-effect comparisons were corrected separately using the Benjamini–Hochberg false discovery rate (FDR). Outcomes used were the mean reaction time (MRT) of the longest delay trial (8 s trials) for the SAT, mean accuracy of the 2-back condition (percentage of correctly identified targets) for the N-back task and two complementary performance measures for the two contrasts of the tracking SST: stop-signal reaction time (SSRT), indexing the speed of response inhibition, and post-stop-error slowing (PES), indexing behavioural adjustment following failed stop trials. PES was included to provide a behavioural measure corresponding specifically to the failed-stop trials examined in the fMRI analyses. SSRT was calculated using the mean method as the MRT on correct go trials minus the mean stop-signal delay and PES was calculated as the MRT on correct go trials immediately following failed stop trials minus the MRT on correct go trials not following a failed stop trial; positive values indicate post-error slowing and negative values indicate post-error speeding. Because all medication conditions for the N-back task deviated from normality (p ≤ 0.05), and given the small sample sizes, within-subject medication effects on all four behavioural performance outcomes in the ADHD group were assessed consistently using Friedman tests and post-hoc Wilcoxon signed-rank tests, whereas ADHD–TD comparisons were assessed using Mann–Whitney U tests. P values for the three post-hoc within- and three post-hoc between-group task comparisons were corrected using FDR. The ADHD sample size for the performance analysis of the SSRT of the tracking SST was smaller (N = 9) than for the brain function effects (N = 12) because three participants who showed negative SSRTs driven by ceiling stop-signal delays (SSD; a high proportion of stops at the maximum SSD) were excluded. For the fMRI activation analyses, these sessions were retained because BOLD contrasts rely on categorical trial types with sufficient trial counts, and bias in SSRT estimation caused by ceiling SSDs does not inherently invalidate event-related BOLD contrasts. For the performance analysis, however, these sessions were excluded because the negative SSRTs would have yielded spurious estimates and inflated the group standard deviations. For the same reason, five participants with negative SSRTs had to be excluded from the TD group, leaving 16 participants in total.

## 3. Results

### 3.1. Demographic data

As expected, baseline Conners 3-P and SCQ scores were higher in participants with ADHD than in TD (both p < 0.001; Table 1). No other group differences were observed (see Table 1).

**Table 1:**
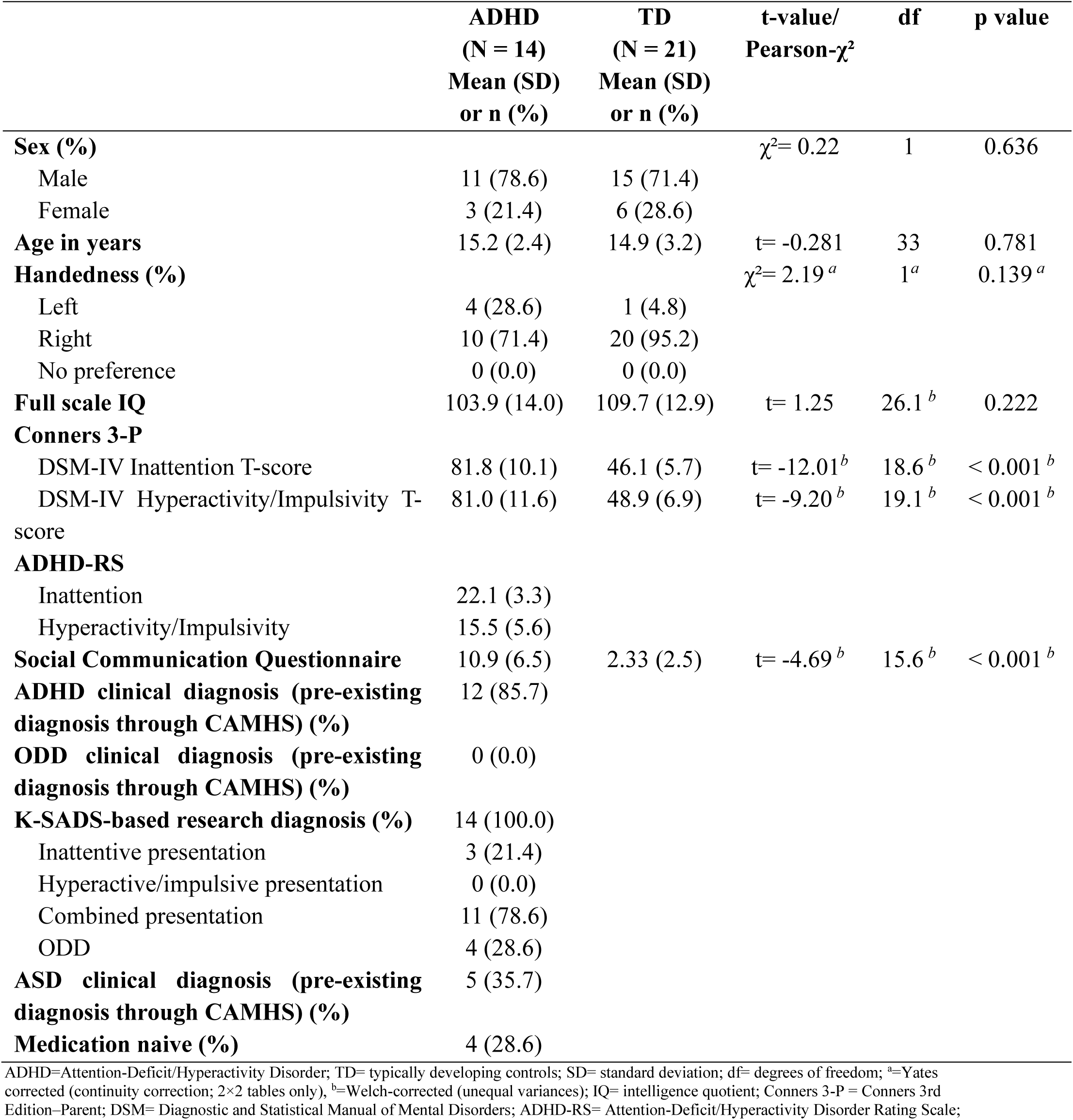
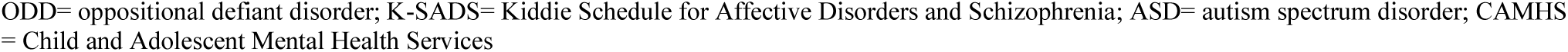
Demographic and clinical data in participants with ADHD and TD.

|  | **ADHD**<br>**(N = 14)**<br>**Mean (SD)**<br>**or n (%)** | **TD**<br>**(N = 21)**<br>**Mean (SD)**<br>**or n (%)** | **t-value/**<br>**Pearson-$\chi^2$** | **df** | **p value** |
| --- | --- | --- | --- | --- | --- |
| **Sex (%)** | | | $\chi^2 = 0.22$ | 1 | 0.636 |
| Male | 11 (78.6) | 15 (71.4) |  |  |  |
| Female | 3 (21.4) | 6 (28.6) |  |  |  |
| **Age in years** | 15.2 (2.4) | 14.9 (3.2) | $t = -0.281$ | 33 | 0.781 |
| **Handedness (%)** | | | $\chi^2 = 2.19^a$ | 1<sup>a</sup> | 0.139<sup>a</sup> |
| Left | 4 (28.6) | 1 (4.8) |  |  |  |
| Right | 10 (71.4) | 20 (95.2) |  |  |  |
| No preference | 0 (0.0) | 0 (0.0) |  |  |  |
| **Full scale IQ** | 103.9 (14.0) | 109.7 (12.9) | $t = 1.25$ | 26.1<sup>b</sup> | 0.222 |
| **Conners 3-P** |  |  |  |  |  |
| DSM-IV Inattention T-score | 81.8 (10.1) | 46.1 (5.7) | $t = -12.01^b$ | 18.6<sup>b</sup> | $< 0.001^b$ |
| DSM-IV Hyperactivity/Impulsivity T-score | 81.0 (11.6) | 48.9 (6.9) | $t = -9.20^b$ | 19.1<sup>b</sup> | $< 0.001^b$ |
| **ADHD-RS** |  |  |  |  |  |
| Inattention | 22.1 (3.3) |  |  |  |  |
| Hyperactivity/Impulsivity | 15.5 (5.6) |  |  |  |  |
| **Social Communication Questionnaire** | 10.9 (6.5) | 2.33 (2.5) | $t = -4.69^b$ | 15.6<sup>b</sup> | $< 0.001^b$ |
| **ADHD clinical diagnosis (pre-existing diagnosis through CAMHS) (%)** | 12 (85.7) |  |  |  |  |
| **ODD clinical diagnosis (pre-existing diagnosis through CAMHS) (%)** | 0 (0.0) |  |  |  |  |
| **K-SADS-based research diagnosis (%)** | 14 (100.0) |  |  |  |  |
| Inattentive presentation | 3 (21.4) |  |  |  |  |
| Hyperactive/impulsive presentation | 0 (0.0) |  |  |  |  |
| Combined presentation | 11 (78.6) |  |  |  |  |
| ODD | 4 (28.6) |  |  |  |  |
| **ASD clinical diagnosis (pre-existing diagnosis through CAMHS) (%)** | 5 (35.7) |  |  |  |  |
| **Medication naive (%)** | 4 (28.6) |  |  |  |  |

ADHD=Attention-Deficit/Hyperactivity Disorder; TD= typically developing controls; SD= standard deviation; df= degrees of freedom; <sup>a</sup>=Yates corrected (continuity correction; 2x2 tables only), <sup>b</sup>=Welch-corrected (unequal variances); IQ= intelligence quotient; Conners 3-P = Conners 3rd Edition-Parent; DSM= Diagnostic and Statistical Manual of Mental Disorders; ADHD-RS= Attention-Deficit/Hyperactivity Disorder Rating Scale;

### 3.2. fMRI drug effects within the ADHD group

#### 3.2.1. ROI analyses

##### 3.2.1.1. Tracking SST

During the SST, there were no significant medication effects in the tested ROIs (p > 0.05).

##### 3.2.1.2. N-back task

During the N-back task (see Table 2 and Figure 2), lisdexamfetamine compared with placebo increased activation in right caudate (T(18) = 4.72, *p*_FWE,SVC_ = 0.011). Both drugs relative to placebo increased activation in left STG (lisdexamfetamine: T(18) = 6.04, *p*_FWE,SVC_ = 0.003; guanfacine: T(18) = 4.94, *p*_FWE,SVC_ = 0.017).

**Figure 2.**
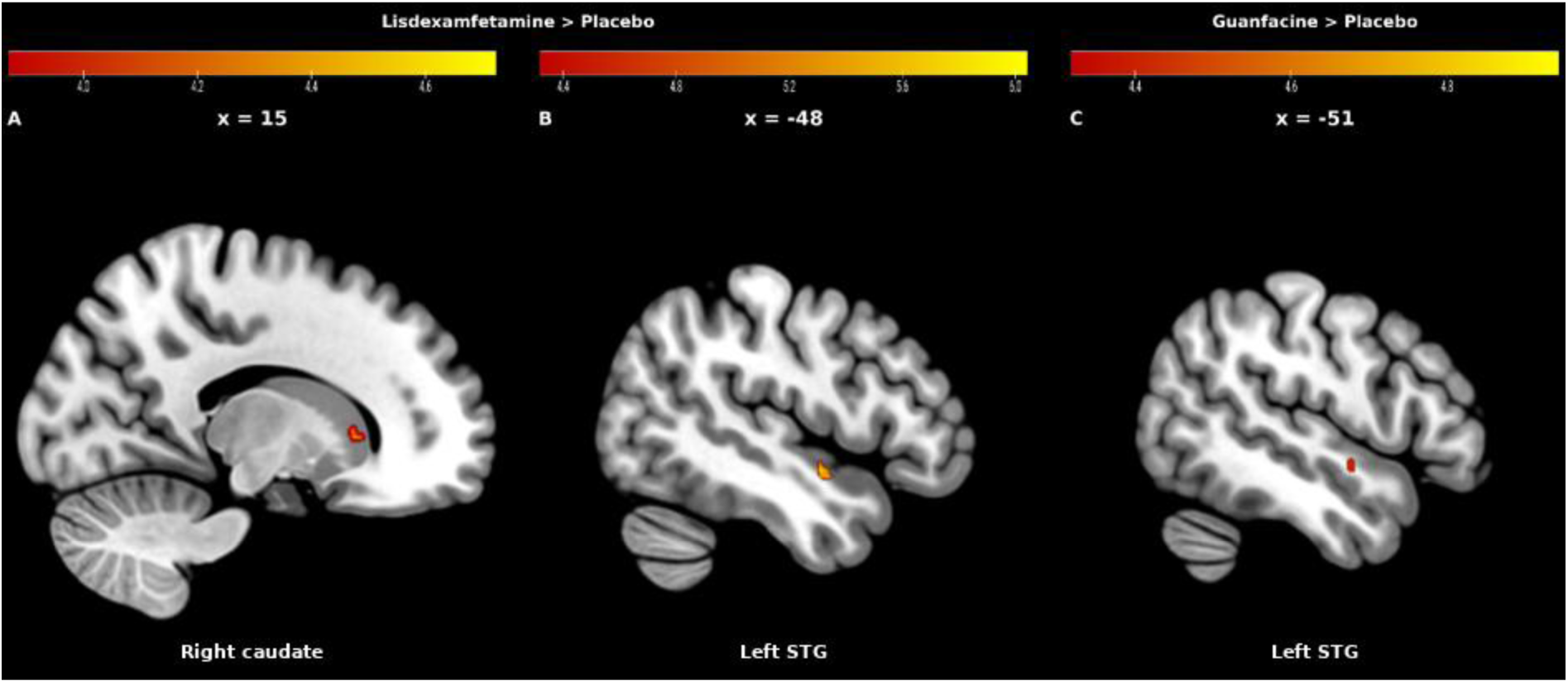
Significant small-volume-corrected activation differences during the N-back task. Sagittal sections are shown at the x-coordinate of the corresponding peak voxel. (A) Lisdexamfetamine > placebo: right caudate (x = 15, y = 20, z = 5; *p*FWE,SVC = 0.011; 15 voxels; critical T = 3.87; observed maximum T = 4.72; surviving voxels T = 3.89–4.72). (B) Lisdexamfetamine > placebo: left superior temporal gyrus (STG; x = −48, y = −4, z = −10; *p*FWE,SVC = 0.003; 25 voxels; critical T = 4.32; observed maximum T = 6.04; surviving voxels T = 4.32–6.04). (C) Guanfacine > placebo: left STG (x = −51, y = −4, z = −7; *p*FWE,SVC = 0.017; 10 voxels; critical T = 4.32; observed maximum T = 4.94; surviving voxels T = 4.40–4.94). Warm colours show the T-statistic maps restricted to the respective a priori ROIs and thresholded at peak-level family-wise error small-volume correction (*p*FWE,SVC < .05). Colour scales are panel-specific and range from the critical threshold to the observed maximum T value. Additional peak-level FWE,SVC-significant ROI findings with k < 10 were observed in the left caudate but are not displayed; see Table 2.

**Table 2:**
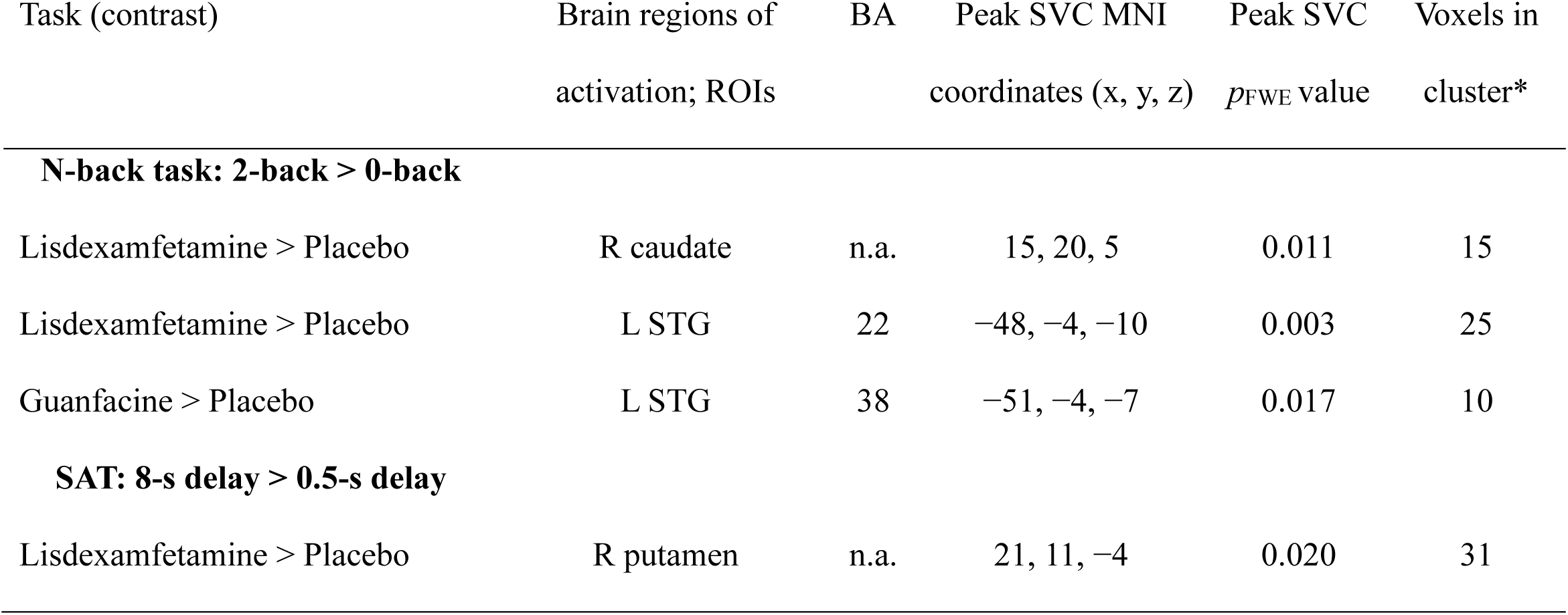
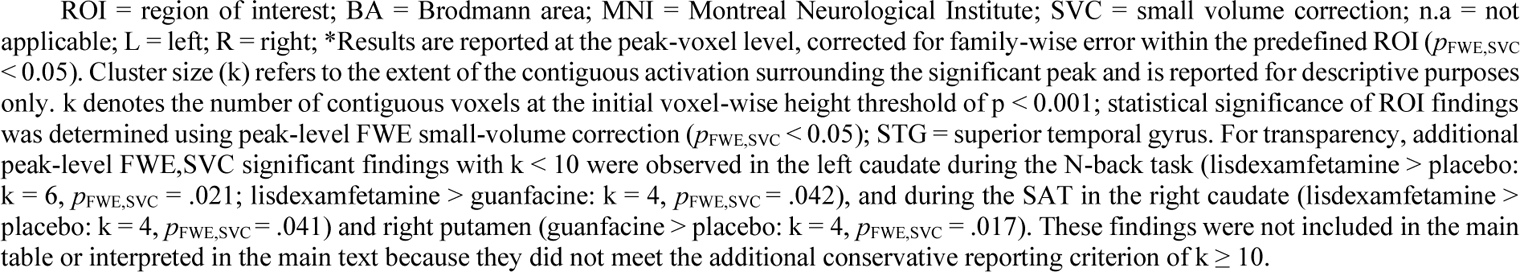
Significant small-volume-corrected ROI findings from pairwise medication contrasts during the N-back task and SAT in participants with ADHD.

| Task (contrast) | Brain regions of activation; ROIs | BA | Peak SVC MNI coordinates (x, y, z) | Peak SVC $p_{FWE}$ value | Voxels in cluster* |
| --- | --- | --- | --- | --- | --- |
| **N-back task: 2-back > 0-back** |  |  |  |  |  |
| Lisdexamfetamine > Placebo | R caudate | n.a. | 15, 20, 5 | 0.011 | 15 |
| Lisdexamfetamine > Placebo | L STG | 22 | -48, -4, -10 | 0.003 | 25 |
| Guanfacine > Placebo | L STG | 38 | -51, -4, -7 | 0.017 | 10 |
| **SAT: 8-s delay > 0.5-s delay** |  |  |  |  |  |
| Lisdexamfetamine > Placebo | R putamen | n.a. | 21, 11, -4 | 0.020 | 31 |

ROI = region of interest; BA = Brodmann area; MNI = Montreal Neurological Institute; SVC = small volume correction; n.a = not applicable; L = left; R = right; \*Results are reported at the peak-voxel level, corrected for family-wise error within the predefined ROI ( $p_{FWE,SVC} < 0.05$ ). Cluster size (k) refers to the extent of the contiguous activation surrounding the significant peak and is reported for descriptive purposes only. k denotes the number of contiguous voxels at the initial voxel-wise height threshold of $p < 0.001$ ; statistical significance of ROI findings was determined using peak-level FWE small-volume correction ( $p_{FWE,SVC} < 0.05$ ); STG = superior temporal gyrus. For transparency, additional peak-level FWE,SVC significant findings with $k < 10$ were observed in the left caudate during the N-back task (lisdexamfetamine > placebo: $k = 6$ , $p_{FWE,SVC} = .021$ ; lisdexamfetamine > guanfacine: $k = 4$ , $p_{FWE,SVC} = .042$ ), and during the SAT in the right caudate (lisdexamfetamine > placebo: $k = 4$ , $p_{FWE,SVC} = .041$ ) and right putamen (guanfacine > placebo: $k = 4$ , $p_{FWE,SVC} = .017$ ). These findings were not included in the main table or interpreted in the main text because they did not meet the additional conservative reporting criterion of $k \geq 10$ .

##### 3.2.1.3. SAT

During the SAT (see Table 2 and Figure 3), lisdexamfetamine compared to placebo increased activation in the right putamen (T(16) = 4.71, *p*_FWE,SVC_ = 0.020).

**Figure 3.**
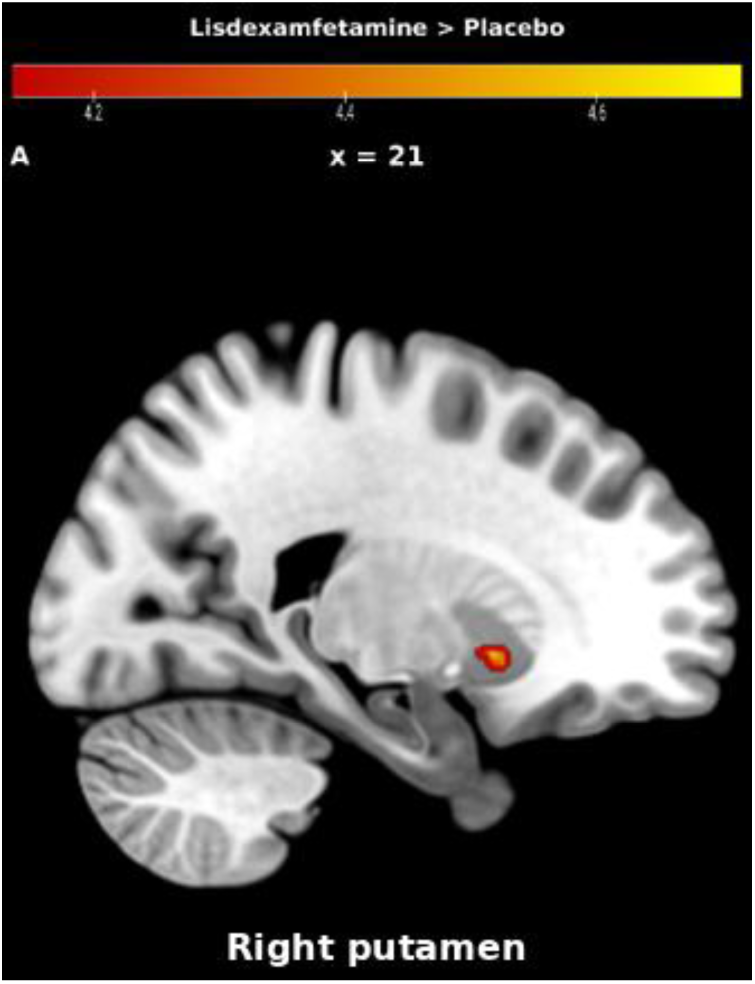
Significant small-volume-corrected activation differences during the SAT. Sagittal sections are shown at the x-coordinate of the corresponding peak voxel. Lisdexamfetamine > placebo: right putamen (x = 21, y = 11, z = −4; *p*FWE,SVC = 0.020; 31 voxels; critical T = 4.14; observed maximum T = 4.71; surviving voxels T = 4.16–4.71). Warm colours show the T-statistic maps restricted to the respective a priori ROI and thresholded at peak-level family-wise error small-volume correction (*p*FWE,SVC < .05). The colour scale ranges from the critical threshold to the observed maximum T value. Additional peak-level FWE,SVC-significant ROI findings with k < 10 were observed in the right caudate and putamen but are not displayed; see Table 2.

#### 3.2.2. Whole-brain analyses

##### 3.2.2.1. Tracking SST

During successful stop vs go trials, the omnibus ANOVA showed a significant cluster in the left/midline precuneus extending to the cuneus (*p*_FWE_ = 0.033, k = 56 voxels; peak MNI coordinates: x = −9, y = −73, z = 38; BA 7), with minor cross-midline extension. A subsequent t-test revealed that lisdexamfetamine compared to guanfacine increased activation in the left precuneus, extending to cuneus, the calcarine cortex and PCC (*p*_FWE_ < 0.001) and right precuneus extending to cuneus and calcarine cortex (*p*_FWE_ = 0.012); see Table 3 and Figure 4.

**Figure 4.**
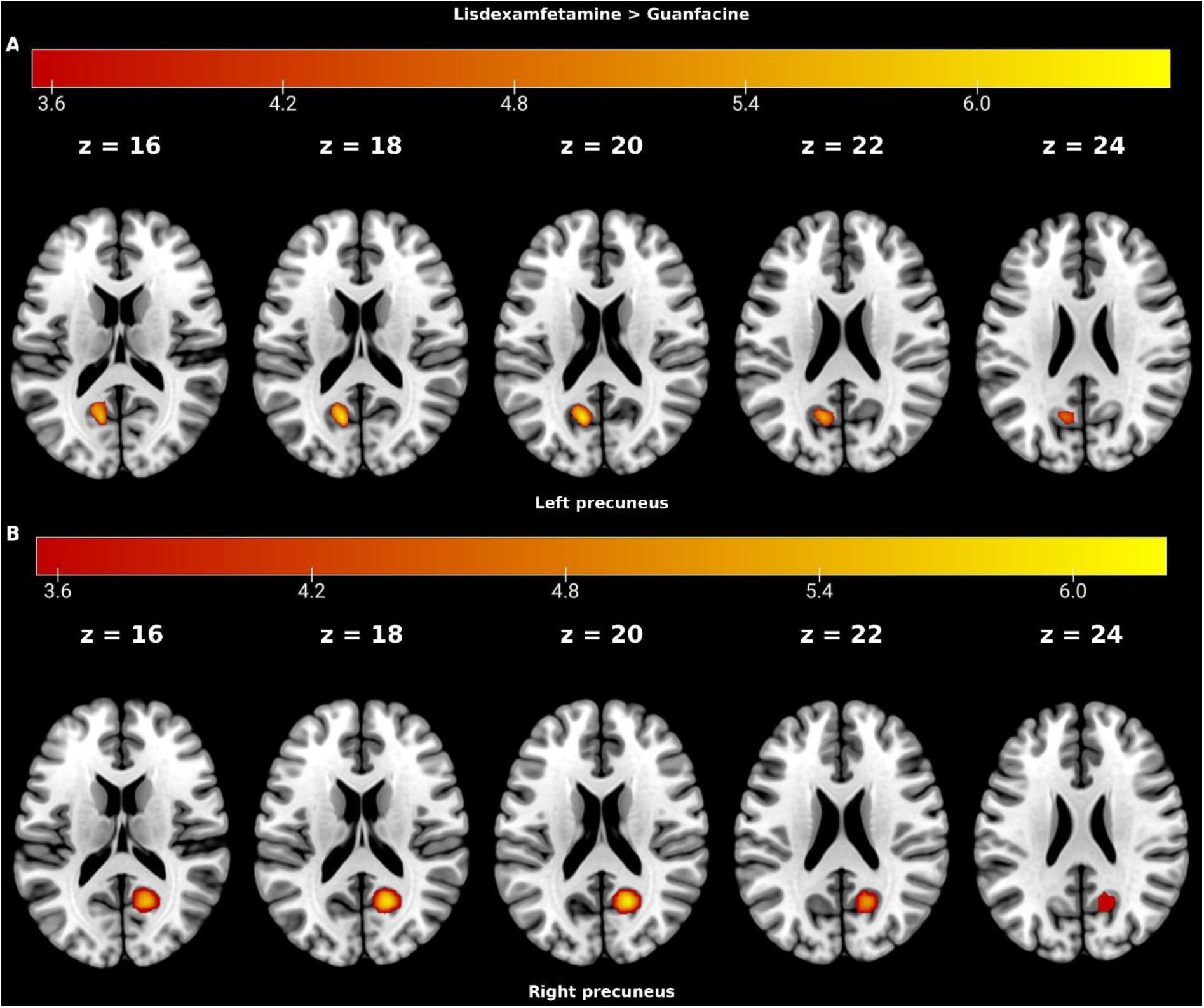
Whole-brain activation differences during successful stop > go trials of the SST. Whole-brain pairwise contrast showing greater activation under lisdexamfetamine than guanfacine in participants with ADHD. (A) Left precuneus cluster extending to cuneus, calcarine cortex and PCC (peak MNI coordinates: x = −12, y = −64, z = 20; cluster-level *p*FWE < 0.001, k = 261). (B) Right precuneus extending to cuneus and calcarine cortex (peak MNI coordinates: x = 18, y = −61, z = 20; cluster-level *p*FWE = 0.012, k = 105). Clusters were identified using an initial voxel-wise threshold of *p*unc < 0.001 (critical T = 3.55) and survived cluster-level family-wise error correction. Warm colours show the voxel-wise T-statistic values within each significant cluster; colour scales range from 3.55 to 6.50 in (A) and from 3.56 to 6.22 in (B). Selected axial sections are shown at the indicated MNI z-coordinates. The left hemisphere is shown on the left.

**Figure 5.**
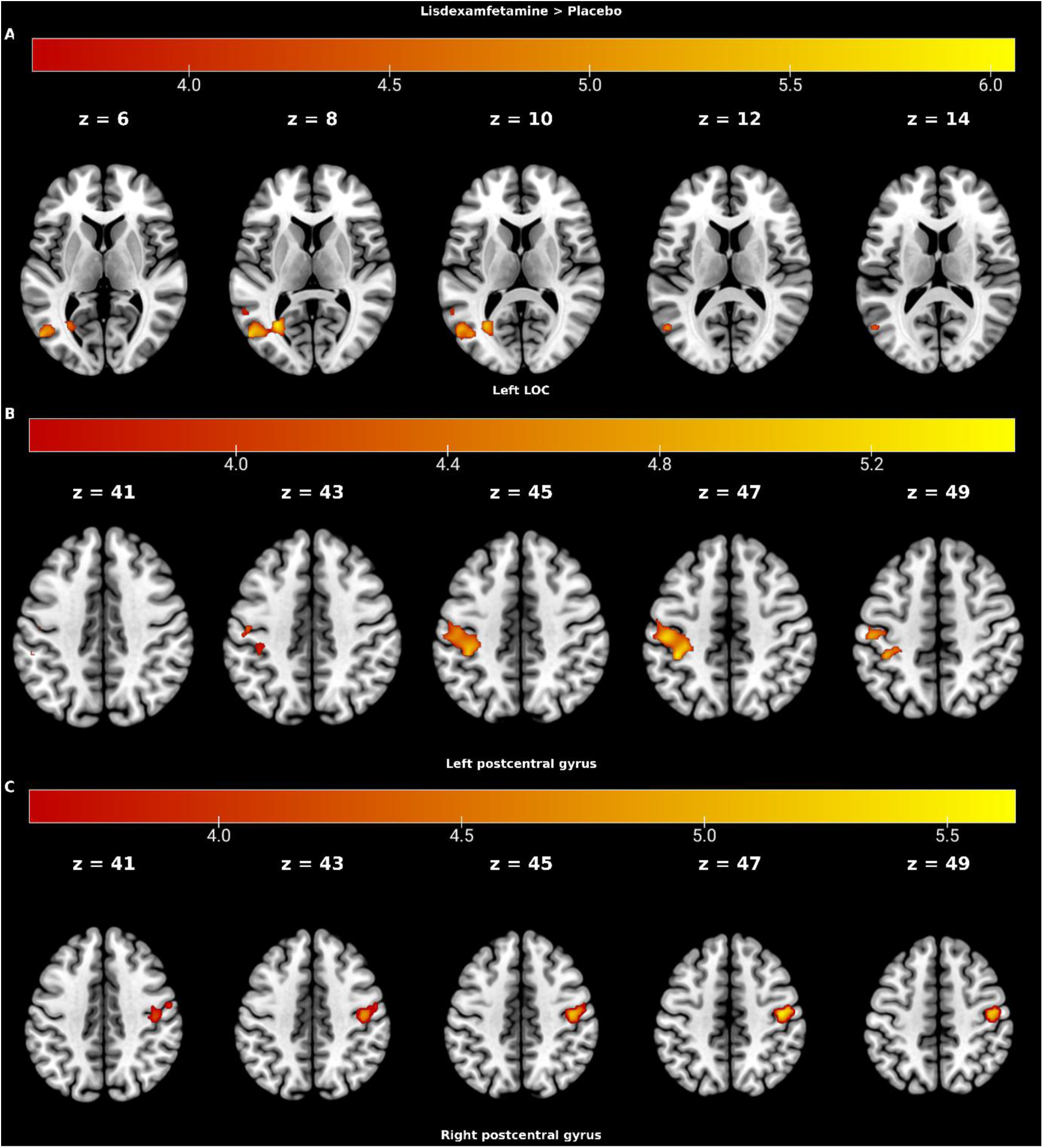
Whole-brain activation differences during the N-back task. Whole-brain pairwise contrast showing greater activation under lisdexamfetamine than placebo in participants with ADHD. (A) Left lateral occipital cortex (LOC) cluster extending to calcarine, MTG, ITG, occipital fusiform, SMG and angular gyrus (peak MNI coordinates: x = −30, y = −64, z = 8; cluster-level *p*FWE = 0.001, k = 200). (B) Left postcentral gyrus cluster extending to precentral gyrus and SPL and SMG (peak MNI coordinates: x = −39, y = −37, z = 47; cluster-level *p*FWE = 0.003, k = 167). (C) Right postcentral gyrus cluster extending to precentral gyrus (peak MNI coordinates: x = 45, y = −16, z = 47; cluster-level *p*FWE = 0.039, k = 85). Clusters were identified using an initial voxel-wise threshold of *p*unc < 0.001 (critical T = 3.61) and survived cluster-level family-wise error correction. Warm colours show the voxel-wise T-statistic values within each significant cluster; colour scales range from 3.61 to 6.07 in (A), from 3.61 to 5.47 in (B), and from 3.62 to 5.65 in (C). Selected axial sections are shown at the indicated MNI z-coordinates. The left hemisphere is shown on the left.

**Table 3:** Significant whole-brain findings from pairwise medication contrasts during the SST and N-back tasks in participants with ADHD.

| Task (contrast) | Brain regions of<br>activation; whole-brain | BA | Peak MNI<br>coordinates (x, y, z) | Cluster<br>$p_{FWE}$ value | Voxels |
| --- | --- | --- | --- | --- | --- |
| **SST: Successful stop vs go trials** |  |  |  |  |  |
| Lisdexamfetamine > Guanfacine | Precuneus/cuneus/<br>L calcarine/PCC | 7 | -12, -64, 20 | < 0.001 | 261 |
|  | R precuneus/cuneus/<br>calcarine | 31 | 18, -61, 20 | 0.012 | 105 |
| **N-back task: 2-back > 0-back** |  |  |  |  |  |
| Lisdexamfetamine > Placebo | L LOC/MTG/ITG/<br>calcarine/occipital<br>fusiform gyrus/SMG/<br>angular gyrus | 19 | -30, -64, 8 | 0.001 | 200 |
| Lisdexamfetamine > Placebo | L postcentral/precentral<br>gyrus/SPL/SMG | 2 | -39, -37, 47 | 0.003 | 167 |
| Lisdexamfetamine > Placebo | R postcentral/precentral<br>gyrus | 3 | 45, -16, 47 | 0.039 | 85 |

BA = Brodmann area; MNI = Montreal Neurological Institute; SST = stop-signal task; L = left; R = right; PCC = posterior cingulate cortex; MTG = middle temporal gyrus; ITG = inferior temporal gyrus; SMG = supramarginal gyrus; SPL = superior parietal lobule; LOC = lateral occipital cortex.

##### 3.2.2.2. N-back task

During 2-back vs 0-back trials of the N-back task, the omnibus ANOVA showed a significant cluster in the left lateral occipital cortex (LOC) extending to the middle temporal gyrus (MTG), angular gyrus and supramarginal gyrus (SMG; *p*_FWE_ = 0.011, k = 85 voxels; peak MNI coordinates: x = −51, y = −67, z = 11; BA 37). A subsequent t-test revealed that lisdexamfetamine compared to placebo increased activation in the left LOC extending to the calcarine cortex, MTG and ITG, occipital fusiform gyrus, SMG and angular gyrus (*p*_FWE_ = .001), left postcentral gyrus extending to the precentral gyrus, SPL and SMG (*p*_FWE_ = .003), and right postcentral gyrus extending to the precentral gyrus (*p*_FWE_ = .039).

##### 3.2.2.3. SAT

During the SAT (8 s vs 0.5 s), the ANOVA showed no significant medication effects in whole-brain analysis (p > 0.05).

### 3.3. Potential normalisation effects of lisdexamfetamine and guanfacine on brain activation differences in ADHD compared to TD

#### 3.3.1. ROI analyses

##### 3.3.1.1. Tracking SST

During successful stop versus go trials of the SST (see Table 4), participants with ADHD under placebo showed lower activation than TD in the left (T(30) = 4.55, *p*_FWE,SVC_ = 0.006) and right putamen (T(30) = 4.04, *p*_FWE,SVC_ = 0.018). Under both medications, these differences were no longer significant. During failed stop versus go trials, participants with ADHD under placebo showed no significant differences compared to TD.

**Table 4:**
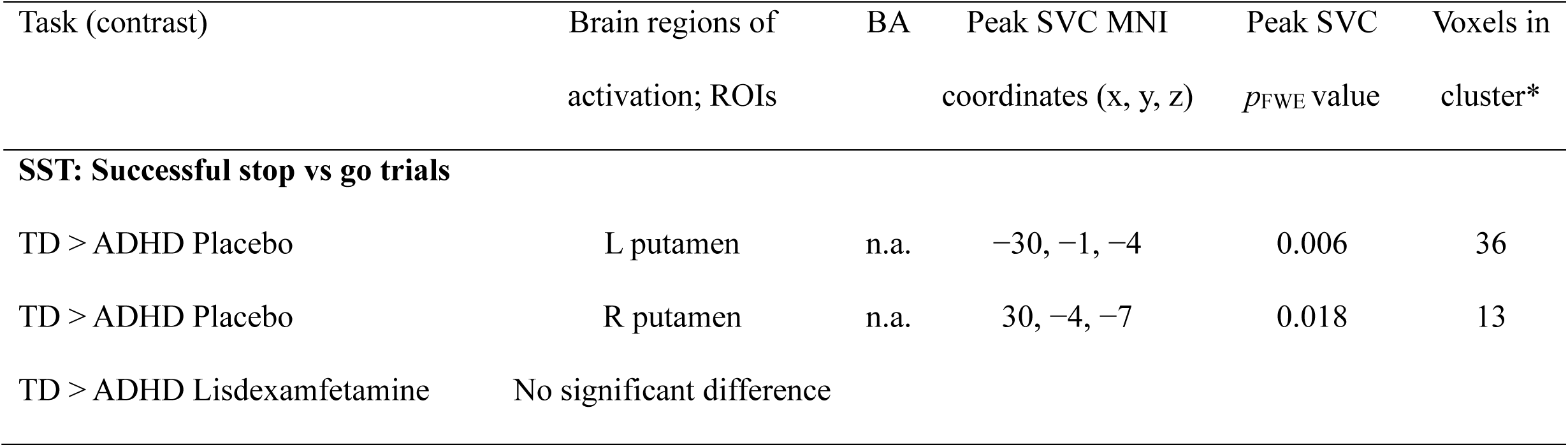
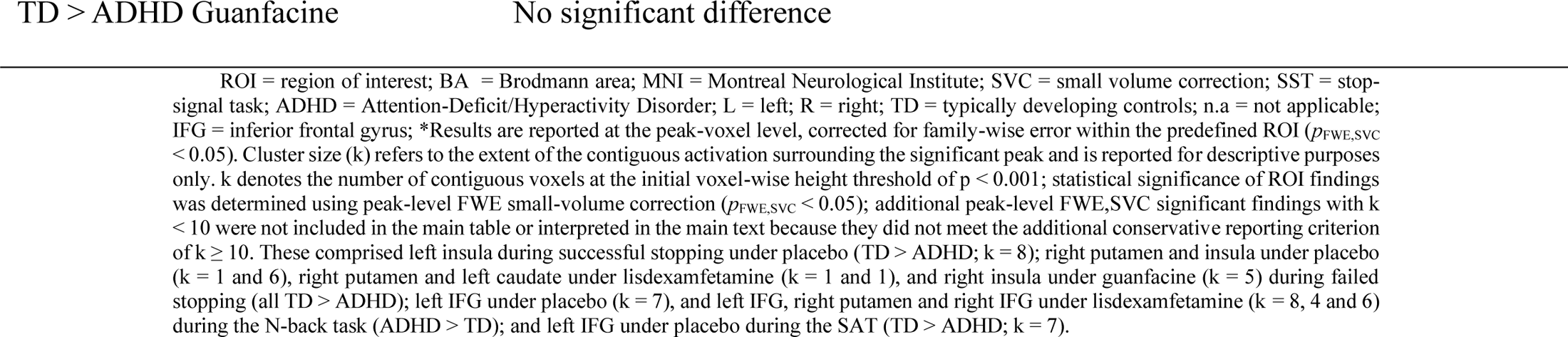
ROI comparisons between TD and ADHD under placebo, lisdexamfetamine and guanfacine during successful stop versus go trials of the SST.

| Task (contrast) | Brain regions of activation; ROIs | BA | Peak SVC MNI coordinates (x, y, z) | Peak SVC $p_{\text{FWE}}$ value | Voxels in cluster* |
| --- | --- | --- | --- | --- | --- |
| **SST: Successful stop vs go trials** |  |  |  |  |  |
| TD > ADHD Placebo | L putamen | n.a. | -30, -1, -4 | 0.006 | 36 |
| TD > ADHD Placebo | R putamen | n.a. | 30, -4, -7 | 0.018 | 13 |
| TD > ADHD Lisdexamfetamine | No significant difference |  |  |  |  |

##### 3.3.1.2. N-back task

During the N-back task, participants with ADHD under placebo showed no significant differences compared to TD.

##### 3.3.1.3. SAT

During SAT, participants with ADHD under placebo showed no significant differences compared to TD.

#### 3.3.2. Whole-brain analyses

##### 3.3.2.1. Tracking SST

During successful stop versus go trials of the SST (see Table 5), participants with ADHD under placebo showed lower activation than TD in the left lingual gyrus extending to the occipital fusiform gyrus, calcarine, occipital pole and right cuneus (*p*_FWE_ < 0.001) and left insula extending to the putamen, pallidum and thalamus (*p*_FWE_ = 0.021). Under lisdexamfetamine, neither of these whole-brain differences remained significant. Under guanfacine, the lower activation in the lingual gyrus remained significant (*p*_FWE_ < 0.001), whereas the left insula difference was no longer significant. In addition, participants with ADHD under guanfacine showed lower activation than TD in the left precuneus extending to the cuneus, PCC and LOC (*p*_FWE_ = 0.024).

**Table 5:** Whole-brain analysis (t-tests) of fMRI comparison between typically developing control subjects (TD) and ADHD during the SST.

| Task (contrast) | Brain regions of<br>activation; whole-brain | BA | Peak MNI<br>coordinates (x, y, z) | Cluster<br>$p_{FWE}$ value | Voxels |
| --- | --- | --- | --- | --- | --- |
| **SST: Successful stop vs go trials** |  |  |  |  |  |
| TD > ADHD Placebo | Lingual gyrus/occipital<br>fusiform gyrus/<br>calcarine/occipital<br>pole/R cuneus | 18 | -6, -73, -4 | < 0.001 | 433 |
| TD > ADHD Placebo | L insula/putamen/<br>pallidum/thalamus | 13 | -33, -22, -1 | 0.021 | 118 |
| TD > ADHD Lisdexamfetamine | No significant<br>difference |  |  |  |  |
| TD > ADHD Guanfacine | Lingual gyrus/<br>calcarine/R cuneus/R<br>occipital fusiform<br>gyrus/ R occipital pole | 18 | 12, -58, 2 | < 0.001 | 283 |
| TD > ADHD Guanfacine | L precuneus/<br>cuneus/PCC/LOC | 31 | -12, -61, 32 | 0.024 | 95 |
| **SST: Failed stop vs go trials** |  |  |  |  |  |
| TD > ADHD Placebo | Calcarine/cuneus/<br>precuneus/lingual<br>gyrus/LOC/R occipital<br>fusiform gyrus/occipital<br>pole | 18 | 6, -79, 17 | < 0.001 | 1061 |
| TD > ADHD Placebo | R parietal operculum/<br>SMG/postcentral gyrus/<br>planum temporale | 40 | 45, -31, 26 | 0.025 | 103 |
| TD > ADHD Lisdexamfetamine | No significant<br>difference |  |  |  |  |
| TD > ADHD Guanfacine | L angular<br>gyrus/MTG/SMG/LOC | 22 | -45, -55, 20 | 0.012 | 123 |
| TD > ADHD Guanfacine | R MTG/LOC/angular<br>gyrus/SMG | 39 | 60, -58, 8 | 0.046 | 81 |

BA = Brodmann area; MNI = Montreal Neurological Institute; SST = stop-signal task; ADHD = Attention-Deficit/Hyperactivity Disorder; TD = typically developing controls; n.a = not applicable; L = left; R = right; PCC = posterior cingulate cortex; LOC = lateral occipital cortex; SMG = supramarginal gyrus; MTG = middle temporal gyrus.

During failed stop versus go trials of the SST, participants with ADHD under placebo showed lower activation than TD in the calcarine extending to the cuneus, precuneus, lingual gyrus, LOC, right occipital fusiform gyrus and occipital pole (*p*_FWE_ < 0.001) and right parietal operculum extending to the SMG, postcentral gyrus and planum temporale (*p*_FWE_ = 0.025). Under lisdexamfetamine, neither of these whole-brain differences remained significant. Under guanfacine, these placebo-related differences were also no longer significant; however, lower activation than TD was observed in the left angular gyrus, extending to the MTG, SMG and LOC (*p*_FWE_ = 0.012) and right MTG extending to the LOC, angular gyrus and SMG (*p*_FWE_ = 0.046).

##### 3.3.2.2. N-back task

During the N-back task there were no significant whole-brain fMRI differences between ADHD on placebo and TD (p > 0.05).

##### 3.3.2.3. SAT

During the SAT there were no significant whole-brain fMRI differences between ADHD on placebo and TD (p > 0.05).

### 3.4. Behavioural task performance within youth with ADHD

Friedman ANOVA showed no within-subject medication effects in the ADHD group for MRT on the SAT (p= 0.067, Kendall’s W = 0.270), accuracy on the N-back task (p= 0.317, Kendall’s W = 0.104), or PES in the SST (p= 0.472, Kendall’s W = 0.063) but showed a significant medication effect on SSRT in the tracking SST (p= 0.032, Kendall’s W = 0.383); see Table 6.

**Table 6:** Behavioural task performance data for youth with ADHD and TD.

| Task Variable | TD<br>N | TD<br>Mean (SD) | ADHD<br>N | ADHD Placebo<br>Mean (SD) | ADHD LDX<br>Mean (SD) | ADHD GFC<br>Mean (SD) |
| --- | --- | --- | --- | --- | --- | --- |
| SST: SSRT (ms) | 16 | 163.2 (89.2) | 9 | 289.3 (65) | 197.7 (97.1) | 264.2 (77.4) |
| PES (ms) | 21 | -70.7 (87.4) | 12 | -79.3 (82.5) | -30.5 (70.1) | -57.4 (80.4) |
| N-back task: accuracy<br>2-back (%) | 20 | 94 (7.5) | 11 | 92.4 (8.70) | 93.9 (5.40) | 87.1 (15.5) |
| SAT: MRT 8 s trials<br>(ms) | 20 | 473.6 (65.3) | 10 | 489.7 (88.1) | 463.7 (61.9) | 517 (89) |

LDX = lisdexamfetamine; GFC = guanfacine; ADHD = Attention-Deficit/Hyperactivity Disorder; TD = typically developing controls; SD = standard deviation; SST = stop-signal task; SSRT = stop-signal reaction time; ms = milliseconds; SAT = sustained attention task; MRT = mean reaction time; PES = post-stop-error slowing

Post-hoc Wilcoxon signed-rank tests revealed that the SSRT in youth with ADHD was significantly lower under lisdexamfetamine compared to placebo (*p*_FDR_ = 0.045, Z = −2.43; Hodges–Lehmann estimated reduction = 93.17 ms, 95% CI [21.22, 167.84]), but not compared to guanfacine (*p*_FDR_ = 0.165, Z = −1.60; estimated reduction = 68.47 ms, 95% CI [−29.16, 168.81]), while SSRT under guanfacine did not differ significantly from SSRT under placebo (*p*_FDR_ = 0.214, Z = −1.24; estimated reduction = 13.88 ms, 95% CI [−31.37, 89.33]).

### 3.5. Behavioural task performance in ADHD vs TD

Participants with ADHD on placebo did not differ from TD in performance in the MRT during the 8 s delay trials of the SAT (*p*_FDR_ = 0.930, Mann–Whitney U = 97, Z = −0.132), accuracy in the N-back task (*p*_FDR_ = 0.697, Mann–Whitney U = 104, Z = −0.495), or PES (*p*_FDR_ = 0.970, Mann–Whitney U = 114, Z= −0.45) in the SST, but differed in the SSRT of the SST (*p*_FDR_ = 0.010, Mann–Whitney U = 20, Z = −2.94; Hodges–Lehmann estimate [TD − ADHD] = −116.30 ms, 95% CI [−192.52, −51.11]). The group difference in SSRT remained significant under guanfacine (*p*_FDR_= 0.019, Mann–Whitney U= 28, Z= −2.49; Hodges–Lehmann estimate = −105.92 ms, 95% CI [−175.70, −23.27]), but not under lisdexamfetamine (*p*_FDR_ = 0.365, Mann– Whitney U = 56, Z = −0.91; Hodges–Lehmann estimate = −30.40 ms, 95% CI [−120.42, 52.34]), compatible with normalisation of the difference.

### 3.6. AEs within youth with ADHD

Friedman ANOVA showed AEs differed across medications (*p* = 0.004, Kendall’s W = 0.390) with post-hoc Wilcoxon signed-rank tests revealing more AEs reported by parents while their children were on lisdexamfetamine compared to placebo (*p*_FDR_ = 0.012, Z = −2.87) and compared to guanfacine (*p*_FDR_ = 0.042, Z= −2.20), whereas guanfacine compared to placebo did not show significantly more AEs (*p*_FDR_ = 0.105, Z = −1.62) (see Supplementary data 5).

## 4. Discussion

This exploratory randomised, placebo-controlled, double-blind, crossover comparative fMRI study examined the comparative effects of lisdexamfetamine, guanfacine and placebo on brain activation and task performance during the SST, N-back task and SAT in youth with ADHD.

ROI analyses showed that during the N-back and SAT, lisdexamfetamine increased right caudate (N-back task) and right putamen (SAT) activation versus placebo while both drugs increased left STG activation during N-back task. In whole-brain analyses during successful inhibition in the SST, lisdexamfetamine increased activation relative to guanfacine in precuneus/cuneus, extending to PCC and calcarine gyrus. During the N-back task, lisdexamfetamine increased similar activation clusters relative to placebo including left occipito-temporo-parietal regions, as well as bilateral postcentral/precentral regions.

For the case-control comparisons, ROI analyses showed that only during successful inhibition in the SST, ADHD vs TD showed reduced putamen activation, which was no longer observed under either drug condition. In whole-brain analyses, placebo-related left insula underactivation during successful stopping was no longer significant under either drug, whereas lingual gyrus underactivation persisted under guanfacine, with additional left precuneus underactivation. During failed stopping, placebo-related calcarine/lingual gyrus and parietal operculum underactivation was no longer significant under either medication, with additional MTG/angular gyrus underactivation under guanfacine relative to TD. Overall, these patterns suggest that both drugs attenuated/possibly normalised several placebo-related case-control activation differences, with lisdexamfetamine having stronger attenuation/possibly normalisation effects (although normalisation was not separately tested) and guanfacine additionally showing reduced MTG/angular gyrus and left precuneus activation.

Lisdexamfetamine also improved and normalised SST performance (SSRT) in youth with ADHD compared to placebo, suggesting performance normalisation.

This is the first study that directly compared the effects of single doses of lisdexamfetamine and guanfacine relative to placebo and each other on task-based fMRI in youth with ADHD and the first study examining the brain function effects of lisdexamfetamine during “cool” executive function tasks. A drug-differential effect was observed during successful inhibition in the SST, where lisdexamfetamine compared to guanfacine showed increased activation effects in precuneus/PCC, key regions for visual-spatial attention and saliency. Youth and adults with ADHD showed precuneus underactivation in a meta-analysis of sustained attention tasks (Hart et al., 2013) and in youth with ADHD during response inhibition (including the PCC) (Rubia et al., 2005). Furthermore, in tasks that demand the allocation of visual-spatial attention to salient stimuli, the precuneus has been previously activated in neurotypical adults during response inhibition (Li et al., 2007; Zhang and Li, 2012). These fMRI findings thus suggest differential effects of these medications on posterior parietal regions of visual-spatial attention, with greater activation under lisdexamfetamine than under guanfacine and oppositely reduced left posterior parietal activation under guanfacine relative to TD.

During the sustained attention and working memory tasks, ROI analyses predominantly showed that lisdexamfetamine increased striatal activation relative to placebo, in right caudate during the N-back task and in right putamen during sustained attention. The increase in striatal activation with lisdexamfetamine during sustained attention is highly novel. For sustained attention and related attention-control tasks, previous whole-brain fMRI studies reported medication-related modulation of inferior frontal, fronto-striato-cerebellar, parieto-temporal, posterior cingulate, and precuneus regions following methylphenidate and atomoxetine (Chou et al., 2015; Kowalczyk et al., 2019; Rubia et al., 2009b). The striatal effects presented here are clinically relevant since meta-analyses in youth with ADHD showed consistent underactivations in basal ganglia including caudate and putamen during tasks of attention (Hart et al., 2013), response inhibition (Lei et al., 2015; Lukito et al., 2020; McCarthy et al., 2014; Norman et al., 2016), and working memory (Wang et al., 2025). These fMRI upregulation effects on basal ganglia are parallel to molecular imaging evidence that stimulant medications strongly engage dopaminergic mechanisms in the basal ganglia. PET studies have shown that stimulant medications increase synaptic dopamine in the striatum (Eklund et al., 2026; Volkow et al., 2002), with a recent meta-analysis showing particularly pronounced psychostimulant-induced dopamine release in the putamen and ventral striatum (Eklund et al., 2026). Furthermore, similarly, direct methylphenidate-related increases in caudate and/or putamen activation have been reported during rewarded attention and response inhibition (Cubillo et al., 2014b; Epstein et al., 2007; Rubia et al., 2009b, 2011, 2014; Vaidya et al., 1998) and basal ganglia underactivation has also been reported to be normalised following methylphenidate during inhibitory-control tasks (Rubia et al., 2011; Shafritz et al., 2004; Vaidya et al., 1998). Last, the meta-analysis of acute stimulant effects in youth with ADHD additionally revealed increased right putamen activation relative to placebo or off medication at a more lenient exploratory threshold (p < .05; Rubia et al., 2014).

To our knowledge, this is the first study to show a direct medication-related increase in STG activation with both lisdexamfetamine and guanfacine during a working-memory task in ADHD. The left STG has been implicated in phonological processing and verbal WM in neurotypical adults (Emch et al., 2019; Turkeltaub and Branch Coslett, 2010). Given that the present N-back task used visually presented letters, the shared increase in left STG activation under lisdexamfetamine and guanfacine may therefore reflect enhanced recruitment of phonological encoding processes supporting verbal working memory. Previous results of studies using the N-back task in fMRI have been inconsistent. While one study in youth with ADHD with single doses of methylphenidate (Kobel et al., 2009), a whole-brain analysis of the same WM sample reported by Cubillo et al. (2014a) (Rubia et al., 2014), and one in adults with 6 weeks of methylphenidate (Biehl et al., 2016) did not find any direct activation effects, the ROI analysis of the same sample showed increased left IFC activation over atomoxetine in an N-back task (Cubillo et al., 2014a) while another study showed left IFC/insula activation compared to no drug in youth with ADHD during another WM task (Prehn-Kristensen et al., 2011). However, lisdexamfetamine showing increased activation in left STG, postcentral gyrus and LOC, during working memory, is similar to findings for methylphenidate in other task domains, e.g. showing increased activation in the left VLPFC/STG relative to placebo and atomoxetine during the SAT (Kowalczyk et al., 2019) and to increased activation in the right occipital lobe compared to placebo in youth with ADHD (Rubia et al., 2011). Right postcentral gyrus activation was further increased under stimulant medication relative to controls across meta-analysed tasks of cognitive control, working memory, attention and reward (Pan et al., 2024). Lisdexamfetamine is a prodrug of dextroamphetamine, and amphetamines have consistently been shown to improve ADHD symptoms via both noradrenergic and dopaminergic mechanisms (Faraone, 2018). The upregulation of these catecholamines may have mediated the medication effects seen in this study. PET studies in neurotypical adults have shown amphetamine-induced dopamine release in striatal, frontal (Narendran et al., 2009; Slifstein et al., 2015), and parietal regions (Narendran et al., 2009).

Lisdexamfetamine had overall stronger and drug-exclusive activation effects on increasing striatal and posterior parieto-occipital areas. This was not expected, because another non-stimulant, atomoxetine, previously normalised left VLPFC underactivation relative to controls and increased right inferior parietal/superior temporal activation relative to placebo during the SST (Cubillo et al., 2014b) and increased right DLPFC activation during WM compared to methylphenidate (Cubillo et al., 2014a). Although alpha-2 agonists like guanfacine differ from noradrenergic reuptake inhibitors like atomoxetine at the molecular level, they nevertheless act on noradrenergic/adrenergic prefrontal mechanisms implicated in arousal, cognition, and ADHD symptom regulation (Neuchat et al., 2023).

The lack of sensitivity for guanfacine effects could be due to the relatively small sample size in our study. Another explanation could be that guanfacine takes longer to show behavioural and neurophysiological effects than lisdexamfetamine (several weeks vs. a few hours) (Mechler et al., 2022); therefore, our design favoured the detection of lisdexamfetamine effects. Nevertheless, the guanfacine-related fMRI effects on the left STG (shared with lisdexamfetamine) are novel, since previous studies examining brain function effects of guanfacine found no direct within-subject activation effects either with a single dose, using fNIRS (Ikeda et al., 2021) or with 6–8 weeks of administration using fMRI (Bédard et al., 2015).

Youth with ADHD on placebo compared to TD showed reduced activation for successful inhibition in the lingual gyrus, putamen and left insula, while for failed stop vs go trials they showed reduced activation in the calcarine gyrus and right parietal operculum. This is similar to meta-analytic findings of underactivation in the putamen during motor and interference inhibition tasks (Lukito et al., 2020; Norman et al., 2016) and parietal lobe during interference inhibition and attention tasks (Hart et al., 2013). The lingual gyrus and cortex surrounding the calcarine sulcus are involved in early visual processing (Palejwala et al., 2021; Vanni et al., 2001) and have previously been shown to be underactivated in youth with ADHD in fMRI studies on response inhibition (Rubia et al., 2005, 2011).

Under lisdexamfetamine, placebo-related case-control activation differences were no longer significant. This pattern is compatible with attenuation/possibly normalisation of placebo-related case-control activation differences under lisdexamfetamine, although this should be interpreted cautiously given the small sample size and the absence of a direct statistical test of normalisation. The attenuation of striato-insular and posterior visual underactivation during the SST extends findings of methylphenidate normalising reduced activation in the insula and right lingual gyrus during the SST (Rubia et al., 2011) and left putamen during timing processing (Rubia et al., 2009a) to another stimulant.

For guanfacine, ROI analyses showed that putamen underactivation was no longer significant, while in whole-brain analyses, lingual gyrus underactivation remained. The absent putamen underactivation under guanfacine is similar to previous findings of the non-stimulant atomoxetine normalising the right putamen underactivation in ADHD relative to TD during the SAT (Kowalczyk et al., 2019).

However, the additional whole-brain underactivation under guanfacine compared to TD observed in the MTG, angular gyrus and left precuneus during the SST could potentially suggest some deactivation or worsening of posterior parietal and temporal activation pattern. Given the involvement of the precuneus in saliency (Rubia et al., 2005; Spinelli et al., 2011) in youth with ADHD and the MTG and broader temporoparietal cortex in stimulus-driven attentional reorienting (Corbetta and Shulman, 2002; Trautwein et al., 2016) in TD, reduced activation in these regions under guanfacine during the SST compared to TD may indicate atypical recruitment of inhibitory and attentional systems in youth with ADHD. This would mirror our neurocognitive data from a larger cohort including the same participants, where guanfacine worsened a performance measure that has been related to attention (RTV) compared to lisdexamfetamine and placebo, while lisdexamfetamine improved MRT and RTV compared to guanfacine and placebo (El Masri et al., 2025). These key areas of saliency and attentional reorienting are furthermore consistently underactivated in youth and adults with ADHD in meta-analyses of task-based fMRI studies, i.e. precuneus (Hart et al., 2013) and right MTG during inhibitory control (Wang et al., 2025) and across different types of tasks (Tamon et al., 2024) in youth with ADHD. This pattern raises the hypothesis that guanfacine may reduce recruitment of posterior attentional and salience-related regions during inhibitory control. An explanation for these potentially underactivating effects on these brain areas with guanfacine is that guanfacine decreases arousal in humans with addiction (Fox et al., 2014), and decreases arousal-related parameters like blood pressure and pulse in youth with ADHD (Kollins et al., 2011). A reduction in arousal could potentially explain the reduced activation, since arousal fluctuations have been shown to modulate thalamo-cortical fMRI signals (Chang et al., 2016), with transient arousal loss during microsleeps linked to reduced thalamic, posterior cingulate, and occipital activity (Poudel et al., 2014). These brain activation mechanisms could potentially shed light on previous findings of no or negative neurocognitive performance effects of guanfacine (Bilder et al., 2016; El Masri et al., 2025; Kollins et al., 2011; Michelini et al., 2023). However, because these underactivation findings derive from comparisons with TD rather than direct guanfacine-versus-placebo reductions, this interpretation remains tentative.

Behaviourally, we found that the performance in the SST (SSRT) in youth with ADHD was significantly improved under lisdexamfetamine, but not under guanfacine, compared to placebo. Lisdexamfetamine also brought values towards TD levels such that the significant ADHD–TD difference observed under placebo in SSRT was no longer significant, suggesting normalisation. This pattern is similar to meta-analyses examining effects of stimulants on cognitive performance in children and adults with ADHD, showing consistent effects on inhibition, reaction time, attention, and WM (Coghill et al., 2014; Isfandnia et al., 2024; Pievsky and McGrath, 2018; Tamminga et al., 2016; Vertessen et al., 2022) and to previous findings showing normalisation of deficient response inhibition under methylphenidate in youth with ADHD (DeVito et al., 2009; Rosch et al., 2016). These behavioural results are also similar to our positive findings in our larger cohort, including the same participants analysed here, of lisdexamfetamine relative to placebo and guanfacine improving MRT and RTV, with the latter being considered an indicator of attention (El Masri et al., 2025). The present SSRT improvement under lisdexamfetamine extends previous reports of improved response inhibition and other cognitive measures following longer-term lisdexamfetamine treatment in youth with ADHD, as well as improved reaction time and vigilance-related measures in adults with ADHD (Coghill et al., 2018; DuPaul et al., 2012; Martin et al., 2014). In contrast, the absence of significant SSRT improvement under guanfacine extends previous studies reporting no improvement in response inhibition, reaction time, sustained attention or working memory following acute or longer-term guanfacine treatment (Bédard et al., 2015; Bilder et al., 2016; El Masri et al., 2025; Kollins et al., 2011). Interestingly, other studies examining single-dose effects of methylphenidate in youth with ADHD during the SST failed to show behavioural improvements (Rubia et al., 2011, 2014). However, this is not uncommon since task-fMRI paradigms are optimised to elicit robust condition-related BOLD activation at the group level and not necessarily elicit performance differences (Elliott et al., 2020). It should be noted that the sample for the ADHD group in the analysis of the SST performance was even smaller than in the fMRI analysis (N = 9 instead of N = 12).

## 5. Limitations

This study has several limitations. First, the sample size of 10–12 youth with ADHD was relatively small for a neuroimaging study, and results hence need replication. However, this is the first study to examine the comparative effects of the more recently licensed ADHD medications lisdexamfetamine and guanfacine on functional brain activation. Second, guanfacine takes longer to show behavioural effects than lisdexamfetamine (several weeks vs a few hours) (Mechler et al., 2022); hence comparative studies of single-dose effects of stimulants and non-stimulants may favour stimulant medication. Third, the age range of participants was relatively broad (8–20 years), spanning substantial developmental changes. Although the within-subject design reduces the risk that age confounded the comparison between treatment conditions, age-related heterogeneity in neural activation and treatment response cannot be ruled out. Fourth, although the ROI analyses applied peak-level small-volume family-wise error correction within each predefined mask, no additional correction for multiple comparisons was applied across the different ROIs. The ROI findings, including the case-control activation differences, should therefore be considered exploratory and require replication in larger samples. Fifth, a substantial proportion of participants showed elevated ASD traits or had an ASD diagnosis, which may have influenced the results since youth and adults with ADHD and ASD have overlapping, but mostly distinctive neural structures and functions (Lukito et al., 2020; Tamon et al., 2024). Co-occurring ASD and ADHD has been associated with distinct neural activation during failed inhibition compared with either disorder alone (Lukito et al., 2023), while methylphenidate-related right prefrontal activation during response inhibition may differ between children with ADHD with versus without co-occurring ASD (Tokuda et al., 2018).

## 6. Conclusion

This is the first study directly comparing the effects of single doses of lisdexamfetamine and guanfacine relative to placebo on task-based fMRI in youth with ADHD. It is also the first study to test lisdexamfetamine fMRI effects on “cool” executive functions in ADHD. Lisdexamfetamine had drug-specific effects of increasing activation in the precuneus/PCC compared to guanfacine during inhibition and drug-exclusive effects relative to placebo in basal ganglia during sustained attention and WM and in posterior parieto-occipital regions during WM. Both drugs shared increased superior temporal activation during WM versus placebo.

Lisdexamfetamine had stronger attenuation/possibly normalisation effects, particularly in the basal ganglia and parieto-occipital regions and further normalised inhibitory performance. These findings are of tentative clinical interest and require replication.

## CRediT Contribution of authorship

All authors listed contributed to the publication: conceptualization (KR), methodology (KR, MAM), funding acquisition (KR, MAM), project administration (SEM, OSK, MC, KR), major parts of participant recruitment and testing (investigation) (SEM, OSK), ad hoc participant recruitment and/or testing (investigation) (MC, SW, SL, SLL, AET, AAC, NB), medical cover or other resources (NM, GSP, VPR, SS, OM, PS, NA, LJ, MR, DO, FA), data analysis (SEM), data curation (SEM), writing the original draft (SEM, KR), edits and reviews (ALL).

## Data Availability

Anonymised participant-level data produced in the present study may be available upon reasonable request to the corresponding authors

Anonymised participant-level data may be made available from the corresponding author upon reasonable request, subject to consistency with the participants’ consent, institutional approval, and an appropriate data-sharing agreement. The data are not publicly available because the consent permits controlled transfer to other researchers but does not explicitly provide for unrestricted public release.

## Acknowledgments

We would like to thank all the participants and their parents for their valuable participation in this study. We would also like to thank the clinical teams and staff at Child and Adolescent Mental Health Services (CAMHS) in South London and Maudsley NHS Foundation Trust, Oxleas NHS Foundation Trust and Great Ormond Street Hospital for Children NHS Foundation Trust (GOSH) for their support with participant recruitment and referrals. We also thank the National Attention Deficit Disorder Information and Support Service (ADDISS), Call for Participants, and the Clinical Record Interactive Search (CRIS), South London and Maudsley NHS Foundation Trust service for supporting participant recruitment and study dissemination. This study represents independent research funded by the National Institute for Health Research (NIHR) Biomedical Research Centre at South London and Maudsley NHS Foundation Trust and King’s College London. The views expressed are those of the author(s) and not necessarily those of the NHS, the NIHR or the Department of Health and Social Care.

## Statements and declarations

### Ethical considerations

The study was approved by the Health Research Authority’s London – London Bridge Research Ethics Committee (reference: 18/LO/0472) and was conducted in accordance with the Declaration of Helsinki.

### Consent to participate

Written informed consent was obtained from participants aged 16 years or older. For participants younger than 16 years, written informed consent was obtained from a parent or legal guardian, and written assent was obtained from the participant.

### Consent for publication

Not applicable.

## Declaration of generative AI and AI-assisted technologies in the writing process

The authors used ChatGPT (OpenAI) to assist with the adaptation and debugging of MATLAB and SPM scripts. This included modifying scripts based on instructions and code provided by the authors, adjusting file paths and parameters, troubleshooting error messages, and implementing code changes. In addition, it was used to support proofreading through author-directed prompts, for example, by identifying grammatical errors, inconsistencies, and unclear wording. All AI-assisted modifications and suggestions were critically reviewed, tested or verified as appropriate, and, where necessary, corrected by the authors. All analytical decisions, interpretations, and final wording were determined by the authors. The authors take full responsibility for the content and accuracy of the published work.

