## Supplementary data for "Differential task-related fMRI effects of a single dose of guanfacine and lisdexamfetamine in youth with ADHD: a randomised, double-blind, placebo-controlled crossover study"

Supplementary data 1: Schematic Fig: sustained attention task (SAT) and full task description

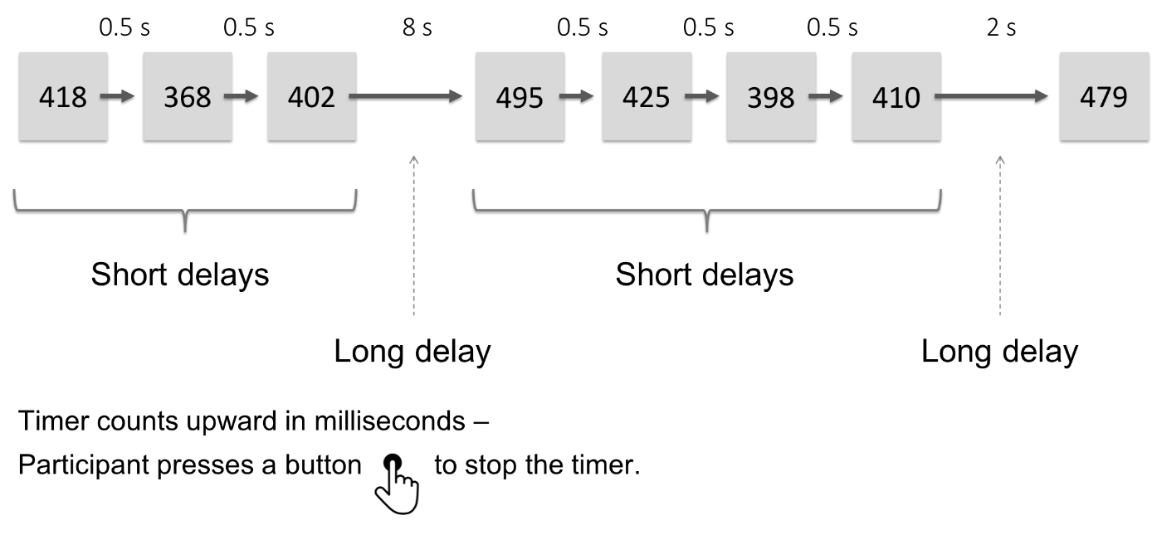

Sustained attention was assessed with a well-validated modified psychomotor vigilance sustained attention fMRI task (SAT) following established protocols (Carlisi et al., 2017; Christakou et al., 2013; Hart et al., 2017; Kowalczyk et al., 2019; Lim et al., 2016; Murphy et al., 2014; Norman et al., 2017). The 12-minute SAT requires participants to respond with a right-handed button press as quickly as possible and within 1 s to a visual stimulus (a timer counting up in milliseconds). The timer appears after either short, predictable, consecutive delays of 0.5 s in series of three to five stimuli (240 trials total) or after unpredictable delays of 2, 5, or 8 s (20 trials each). Trials involving longer delays were pseudorandomly interspersed into the 0.5 s series after at least three 0.5 s trials. The long, infrequent, unpredictable delays require a greater sustained attention load which increases with increasing length of delay, while the short, predictable 0.5 s trials are typically anticipated and therefore place a higher demand on sensorimotor synchronisation (Christakou et al., 2013; Murphy et al., 2014).

Supplementary data 2: Schematic figure of the tracking stop-signal task (SST) and full task description

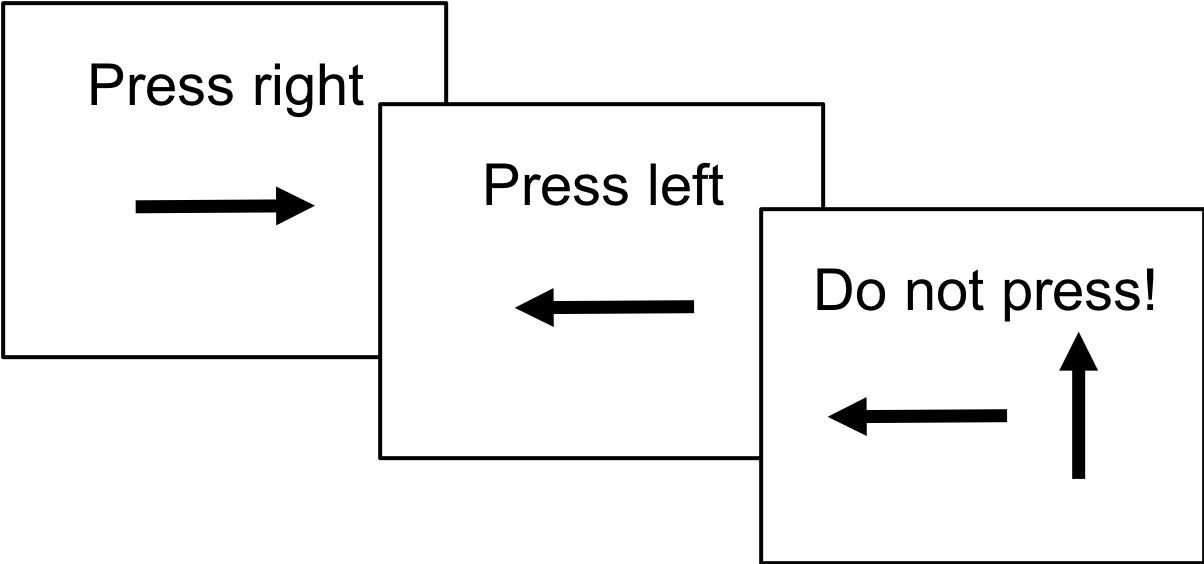

Response inhibition was measured with an established, well-validated, mixed-trials, event-related, individually adjusted tracking SST (Costa et al., 2013; Cubillo et al., 2014b; Hart et al., 2014; Rubia et al., 1999, 2003, 2005, 2007, 2008, 2010, 2013). Participants practised the 6-min mixed-trials, event-related tracking SST once, which measures the ability to suppress an already triggered motor response. The basic go task is a choice reaction time task with a mean intertrial interval of 1.8 s, where participants have to respond to go arrows of 500 ms duration (≈80% of trials, 156 trials) pointing either right or left with a right or left button response with the right/left thumb, followed by a blank of 1300 ms. In ≈20% of trials (40 trials), the go-signals are followed and sometimes interrupted (about 250 ms later) by stop-signals, and participants have to inhibit their motor responses. A tracking algorithm changes the time interval between go-signal and stop-signal onsets according to each subject’s performance on previous trials based on the average percentage of inhibition over previous stop trials, recalculated after each stop trial, resulting in 50% successful and 50% unsuccessful inhibition trials.

Supplementary data 3: Schematic figure of the N-back working memory task (N-back task) and full task description

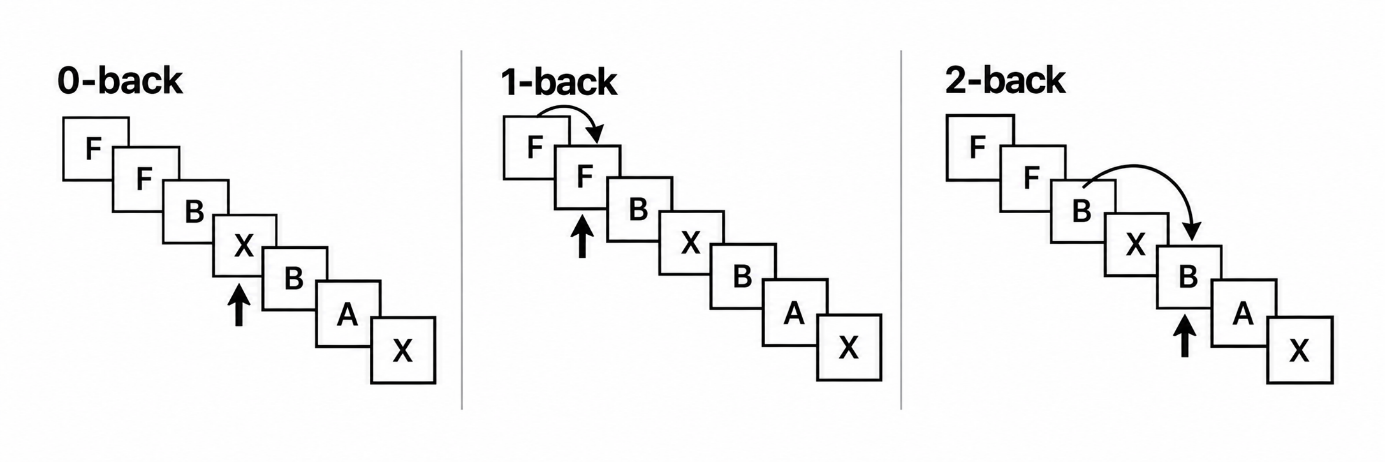

Arrow points to the target

The N-back task was adapted from previous block-design paradigms to include only 0-back, 1-back, and 2-back conditions (Chantiluke et al., 2015; Cubillo et al., 2014a; Kobel et al., 2009), excluding the 3-back to ensure a suitable level of difficulty for younger children. Subjects practised the task once before scanning. The 6 min 12 s block-design WM task consists of three conditions. During ‘1-back’ and ‘2-back’ conditions, subjects are presented with a series of letters (A–Z) with stimulus onsets 2 s apart and must respond with their right thumb using a button box whenever the letter presented is the same as one or two trials before it, respectively (e.g. 2-back: B/J/A/J). This requires both storage and continuous updating of stimuli being held in WM. In the baseline vigilance ‘0-back’ condition, subjects must respond to each X that appears on the screen. The task consists of 12 randomised blocks. Before each block, written instructions of 3 s duration are shown as to which condition is next (i.e. ‘0-back’, ‘1-back’, etc.). Each block contains 14 stimuli, including three targets and 11 non-targets. Each condition is presented four times.

Supplementary data 4: fMRI acquisition parameters for all three tasks

**SAT:** T2*-weighted gradient-echo EPI images depicting BOLD contrast were acquired (TR = 1.5 s, TE = 30 ms, flip angle = 70°, in-plane voxel size = 3.75 × 3.75 mm, slice thickness = 3.5 mm, slice gap = 1.0 mm; 31 axial slices; matrix 64 × 64; 480 volumes).

**SST:** T2*-weighted gradient-echo EPI BOLD images were acquired (TR = 1.8 s, TE = 30 ms, flip angle = 75°, in-plane voxel size = 3.30 × 3.30 mm, slice thickness = 3.0 mm, slice gap = 0.3 mm; matrix 64 × 64; 38 slices; 200 volumes).

**N-back task:** T2*-weighted gradient-echo EPI BOLD images were acquired (TR = 2.0 s, TE = 30 ms, flip angle = 75°, with an in-plane voxel size of 3.75 × 3.75 mm, slice thickness of 3.0 mm (gap 0.3 mm), a matrix 64 × 64, 41 slices and 186 volumes).

Supplementary data 5: Reported adverse effects

| **Reported adverse effect (%)** | **Lisdexamfetamine (N = 14)** | **Guanfacine (N = 14)** | **Placebo (N = 14)** |
| --- | --- | --- | --- |
| Fatigued | 6 (42.9%) | 4 (28.6%) | 4 (28.6%) |
| Poor appetite | 10 (71.4%) | 2 (14.3%) | 2 (14.3%) |
| Drowsy | 4 (28.6%) | 4 (28.6%) | 3 (21.4%) |
| Has trouble getting off to sleep | 7 (50.0%) | 0 (0.0%) | 1 (7.1%) |
| Irritable mood | 4 (28.6%) | 1 (7.1%) | 2 (14.3%) |
| Complains of headache | 3 (21.4%) | 1 (7.1%) | 2 (14.3%) |
| Complains of stomachache | 5 (35.7%) | 3 (21.4%) | 1 (7.1%) |
| Excited | 4 (28.6%) | 1 (7.1%) | 2 (14.3%) |
| Stares a lot or daydreams | 3 (21.4%) | 2 (14.3%) | 1 (7.1%) |
| Easily distracted | 3 (21.4%) | 3 (21.4%) | 2 (14.3%) |
| Talks less than usual with other children/peers | 4 (28.6%) | 2 (14.3%) | 0 (0.0%) |
| Seems unsteady | 2 (14.3%) | 1 (7.1%) | 1 (7.1%) |
| Dizziness | 3 (21.4%) | 1 (7.1%) | 0 (0.0%) |
| Looks sad, miserable | 1 (7.1%) | 0 (0.0%) | 0 (0.0%) |
| Angry | 0 (0.0%) | 0 (0.0%) | 0 (0.0%) |
| Flushing | 1 (7.1%) | 0 (0.0%) | 0 (0.0%) |
| Not interested in other children/peers | 3 (21.4%) | 1 (7.1%) | 1 (7.1%) |
| Complains about light headedness and muscular weakness | 1 (7.1%) | 0 (0.0%) | 0 (0.0%) |
| Complains about dry mouth | 2 (14.3%) | 1 (7.1%) | 1 (7.1%) |
| Complains about palpitation | 2 (14.3%) | 1 (7.1%) | 0 (0.0%) |
| Looks anxious | 3 (21.4%) | 0 (0.0%) | 0 (0.0%) |
| Low confidence | 3 (21.4%) | 0 (0.0%) | 0 (0.0%) |
| Displays twitches (tics) | 2 (14.3%) | 0 (0.0%) | 0 (0.0%) |
| Complains about constipation | 0 (0.0%) | 0 (0.0%) | 0 (0.0%) |
| Crying spells | 0 (0.0%) | 0 (0.0%) | 0 (0.0%) |
| Has nightmares | 0 (0.0%) | 0 (0.0%) | 0 (0.0%) |
| Scratches himself/herself more, bites his/her nails or his/her lips more | 1 (7.1%) | 0 (0.0%) | 1 (7.1%) |

Supplementary data 6: CONSORT reporting checklist (extension to randomized crossover trials)

|  |  | **Reporting Item** | **Page number** |
| --- | --- | --- | --- |
| **Title and Abstract** |  |  |  |
| Title† | #1a | Identification as a randomised crossover trial in the title | 1 |
| Abstract† | #1b | Specify a crossover design and report all information outlined in table 2 | 3 |
| **Introduction** |  |  |  |
| Background‡ | #2a | Scientific background and explanation of rationale | 5–8 |
| Objectives‡ | #2b | Specific objectives or hypotheses | 8-9 |
| **Methods** |  |  |  |
| Trial design† | #3a | Rationale for a crossover design. Description of the design features including allocation ratio, especially the number and duration of periods, duration of washout period, and consideration of carry over effect | 9–13 |
| Change from protocol‡ | #3b | Important changes to methods after trial commencement (such as eligibility criteria), with reasons | 13–14 |
| Participants‡ | #4a | Eligibility criteria for participants | 10–12 |
| Settings and location‡ | #4b | Settings and locations where the data were collected | 10, 15 |
| Interventions† | #5 | The interventions with sufficient details to allow replication, including how and when they were actually administered | 13–14 |
| Outcomes‡ | #6a | Completely defined prespecified primary and secondary outcome measures, including how and when they were assessed | 14-19 |
| Changes to outcomes‡ | #6b | Any changes to trial outcomes after the trial commenced, with reasons | n/a |
| Sample size† | #7a | How sample size was determined, accounting for within participant variability | 9–10 |
| Interim analyses and stopping guidelines‡ | #7b | When applicable, explanation of any interim analyses and stopping guidelines | n/a |
| **Randomisation** |  |  |  |
| Sequence generation‡ | #8a | Method used to generate the random allocation sequence | 13 |
| Sequence generation‡ | #8b | Type of randomisation; details of any restriction (such as blocking and block size) | 13 |
| Allocation concealment mechanism‡ | #9 | Mechanism used to implement the random allocation sequence (such as sequentially numbered containers), describing any steps taken to conceal the sequence until interventions were assigned | 13 |
| Implementation† | #10 | Who generated the random allocation sequence§, who enrolled participants, and who assigned participants to the sequence of interventions | 13 |
| Blinding‡ | #11a | If done, who was blinded after assignment to interventions (for example, participants, care providers, those assessing outcomes) and how | 13 |
| Similarity of interventions‡ | #11b | If relevant, description of the similarity of interventions | 13 |
| Statistical methods† | #12a | Statistical methods used to compare groups for primary and secondary outcomes which are appropriate for crossover design (that is, based on within participant comparison)¶ | 16–19 |
| Additional analyses‡ | #12b | Methods for additional analyses, such as subgroup analyses and adjusted analyses | 17–19 |
| **Results** |  |  |  |
| Participant flow (a diagram is strongly recommended)† | #13a | The numbers of participants who were randomly assigned, received intended treatment, and were analysed for the primary outcome, separately for each sequence and period | 20 |
| Losses and exclusions† | #13b | Number of participants excluded at each stage, with reasons, separately for each sequence and period | 10–12, 20  Partially reported: losses and exclusions with reasons are reported overall in the Participants section (pp. 10–12) and Figure 1 (p. 20), but not separately by randomised treatment sequence and period. |
| Recruitment‡ | #14a | Dates defining the periods of recruitment and follow-up | 10 |
| Trial end‡ | #14b | Why the trial ended or was stopped | n/a |
| Baseline data† | #15 | A table showing baseline demographic and clinical characteristics by sequence and period | 21  Baseline demographic and clinical characteristics are reported overall in Table 1 (p. 21); these are not presented separately by randomised treatment sequence. Period-specific baseline outcome measures were not reported. |
| Numbers analysed† | #16 | Number of participants (denominator) included in each analysis and whether the analysis was by original assigned groups | 10–12; 18–19; 21; 32-33 |
| Outcomes and estimation† | #17a | For each primary and secondary outcome, results including estimated effect size and its precision (such as 95% confidence interval) should be based on within participant comparisons. In addition, results for each intervention in each period are recommended | 22–33  Hodges–Lehmann estimates with 95% confidence intervals are reported for pairwise within-participant behavioural comparisons. Neuroimaging results are reported using peak-level test statistics and corrected significance values. |
| Binary outcomes‡ | #17b | For binary outcomes, presentation of both absolute and relative effect sizes is recommended | n/a – no binary outcomes |
| Ancillary analyses‡ | #18 | Results of any other analyses performed, including subgroup analyses and adjusted analyses, distinguishing prespecified from exploratory | 22–33 |
| Harms† | #19 | Describe all important harms or unintended effects in a way that accounts for the design (for specific guidance, see CONSORT for harms) | 13–14, 33, 57–58 |
| **Discussion** |  |  |  |
| Limitations† | #20 | Trial limitations, addressing sources of potential bias, imprecision, and if relevant, multiplicity of analyses. Consider potential carry over effects | 40–41 |
| Generalisability‡ | #21 | Generalisability (external validity, applicability) of the trial findings | 40–41 |
| Interpretation‡ | #22 | Interpretation consistent with results, balancing benefits and harms, and considering other relevant evidence | 33–42 |
| **Other information** |  |  |  |
| Registration‡ | #23 | Registration number and name of trial registry | 4, 9 |
| Protocol‡ | #24 | Where the full trial protocol can be accessed, if available | 4, 9 |
| Funding‡ | #25 | Sources of funding and other support (such as supply of drugs), role of funders | 43-44 |

† Modified original CONSORT item.

‡ Unmodified CONSORT item.

§ Random sequence refers to a list of random orders, typically computer generated; it should not be confused with the sequence of interventions received by an individual participant.

¶ A within participant comparison accounts for the correlation between measurements because each participant acts as their own control.
